# Comparison of ChatGPT and Gemini as sources of references in otorhinolaryngology

**DOI:** 10.1101/2024.08.12.24311896

**Authors:** W. Wiktor Jędrzejczak, Małgorzata Pastucha, Henryk Skarżyński, Krzysztof Kochanek

## Abstract

**Introduction:** An effective way of testing chatbots is to ask them for references since such items can be easily verified. The purpose of this study was to compare the ability of ChatGPT-4 and Gemini Advanced to select accurate references on common topics in otorhinolaryngology.

**Methods:** ChatGPT-4 and Gemini Advanced were asked to provide references on 25 topics within the otorhinolaryngology category of Web of Science. Within each topic, we set as target the most cited papers which had “guidelines” in the title. The chatbot responses were collected on three consecutive days to take into account possible variability. The accuracy and reliability of the provided references were evaluated.

**Results:** Across the three days, the accuracy of ChatGPT-4 was 29–45% while that of Gemini Advanced was 10–17%. Common errors included false author names, false DOI numbers, and incomplete information. Lower percentage errors were associated with higher number of citations.

**Conclusions:** Both chatbots performed poorly in finding references, although ChatGPT-4 provided higher accuracy than Gemini Advanced.

## Introduction

Chatbots based on large language models (LLMs) are increasingly being tested across various domains for their ability to provide accurate and reliable information [1]. However, the field of otorhinolaryngology, which deals with disorders of the ear, nose, and throat (ENT), presents a unique challenge to these chatbots due to the specialized and often complex nature of its scientific literature [2]. One method to evaluate the accuracy of a chatbot is to examine the references to scientific papers provided in response to a user query. This approach not only tests the chatbot’s ability to access and retrieve the relevant literature but also its ability to discern the most credible sources.

The debate on the accuracy and reliability of chatbots based on LLMs is ongoing. However, their performance can be verified and quantified more easily when they are asked to provide references rather than open-ended responses, which are more subjective and harder to validate. Typically, LLMs are trained on extensive datasets, and it is therefore likely that highly cited knowledge will be more accessible and more readily retrieved. However, research so far into the references provided by chatbots has found a peculiar problem – the fabrication of references [3–5].

Some of the most well-known chatbots based on LLM are OpenAI’s ChatGPT and Google’s Gemini. Recent studies have specifically evaluated ChatGPT’s ability to provide references in the field of otorhinolaryngology [6, 7]. One report indicates that ChatGPT can achieve up to 87% accuracy in delivering appropriate references [7]. However, the cited studies also highlight significant errors, which raises questions about the consistency and reliability of the information chatbots provide. It is well-documented that ChatGPT-4 offers improved performance over its predecessor, ChatGPT-3.5, but the performance of other models, such as Gemini, remains largely unexplored.

A study conducted about a year ago using earlier versions of chatbots, specifically ChatGPT-3.5 and Bard (the precursor to Gemini), revealed severe limitations in their ability to provide accurate references [8]. Given the rapid advancements in LLMs, it is of interest to reassess the capabilities of the latest versions.

This study aims to compare the accuracy of references provided by the most advanced versions of ChatGPT and Gemini. By systematically evaluating and comparing their performance in the context of otorhinolaryngology, this research seeks to identify which model currently offers better accuracy and reliability in referencing the scientific literature in this specialized field. Such an assessment might help us understand the potential and limitations of chatbots in supporting professionals and researchers within otorhinolaryngology.

## Method

Two chatbots based on LLMs were tested: ChatGPT-4 (Open AI, USA) and Gemini Advanced (Google, USA). We based our research on scientific articles that are guidelines on various topics in otorhinolaryngology. We assumed that topics related to the guidelines would be widely covered in the training space used for chatbots, since they can be referred to not only in other scientific articles, but also in books and websites. As such, it can be expected that chatbots will have access to this information.

The topics were selected as follows. The Web of Science was searched for papers with “guideline” in the title. Then the search was limited to the otorhinolaryngology category on Web of Science. Repeating topics were removed (e.g. papers which had the same title but with “update” added). Papers with at least 100 citations were then selected. This resulted in 25 papers which formed the basis of a list of topics, as shown in Table 1.

**Table 1.**
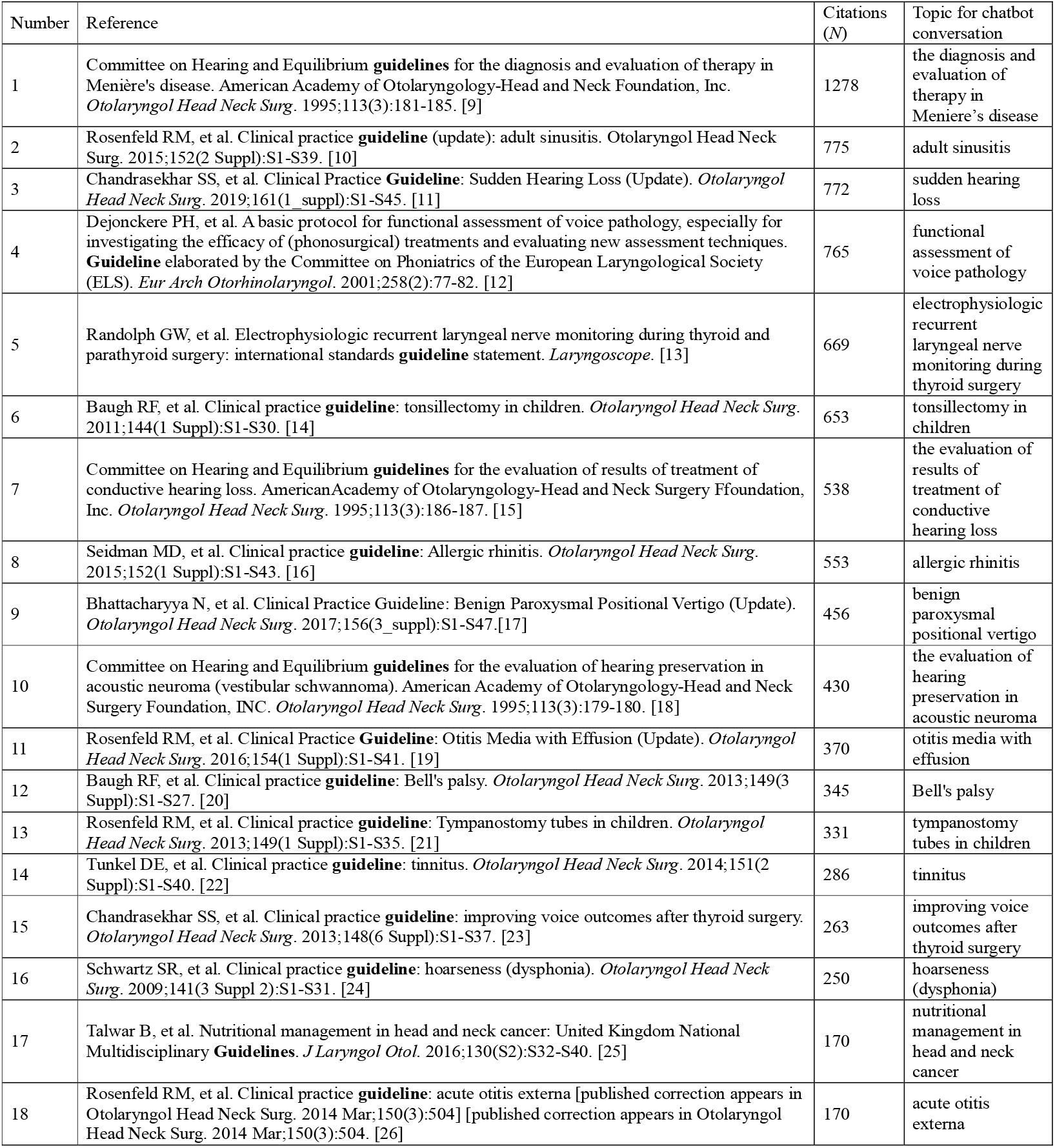
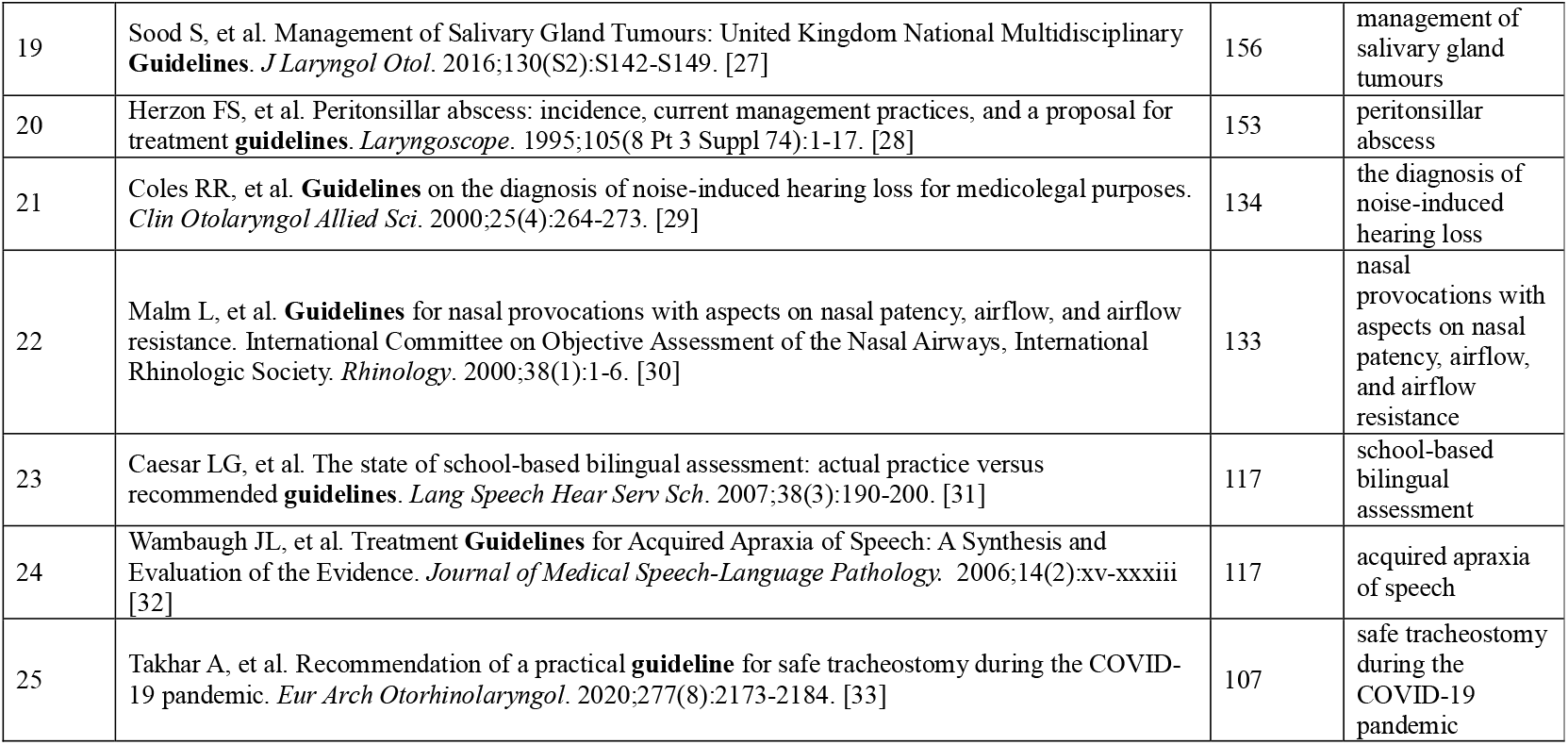
The highly cited publications, with “guideline” in the title, which served as targets. The number of citations they have received in Web of Science is listed. From these papers, a list of chat “topics” was created, which were then used for framing queries to the chatbots.

The prompt for the chatbots was as follows:

“Please provide references to scientific papers on guidelines for [‘topic’]. Only the bibliographic data of the papers is required.”

The prompts were entered separately and the chatbots were reset to a new conversation after each question. The responses of ChatGPT-4 and Gemini Advanced were collected on three consecutive days (8–10.07.2024). The references found by the chatbots (see supplementary file) were verified with Pubmed, Web of Science, and Google Scholar. We checked whether: all were correct; all were accurate except that the Digital Object Identifier (DOI) was not given; all were accurate but the wrong DOI number was given; partially accurate but missing some information (but no false information); partially accurate but with some false information; and totally false information.

Both tested chatbots often added links to certain webpages in their responses. This was not analyzed since the questions explicitly asked for references, and any additional information provided by the chatbots was omitted.

### Statistical methods

All analyses were made in Matlab (version 2023b, MathWorks, Natick, MA). Fleiss Kappa was used to evaluate consistency [34]. The values of Kappa can be interpreted as <0.0, poor; 0.01–0.2, slight; 0.21–0.4, fair; 0.41–0.6, moderate; 0.61–0.8, substantial; and 0.81–1.0, almost perfect agreement [35]. Chi-squared tests were used to assess differences. In all analyses, a 95% confidence level (*p* < 0.05) was taken as the criterion of significance.

## Results

In the responses to each of the 25 questions framed in terms of the ‘topics’ in Table 1, chatbots usually provided more than one reference. Table 2 shows the accuracy of these references across the three sessions as retrieved by ChatGPT-4 and Gemini Advanced. By accuracy we mean only the correctness of the reference(s) provided. Table 2 shows the numbers and percent accuracy; it also divides inaccurate references into subgroups showing the nature of the errors. For ChatGPT the number of accurate references varied from 29% to 45% across three sessions, while for Gemini it varied from 10% to 17%. ChatGPT was significantly better than Gemini for all sessions. As accurate references we included those that had all the correct information except a missing DOI.

**Table 2.** General accuracy of ChatGPT-4 and Gemini Advanced across three sessions. The number of references found is given together with the percentage in parentheses. The accuracy for each session (ChatGPT-4 vs Gemini Advanced) were compared using Chi-squared tests, with asterisks showing statistical significance.

|  | ChatGPT-4 |  |  | Gemini Advanced |  |  |
| --- | --- | --- | --- | --- | --- | --- |
|  | Session 1,<br>N (%) | Session 2,<br>N (%) | Session 3,<br>N (%) | Session 1,<br>N (%) | Session 2,<br>N (%) | Session 3,<br>N (%) |
| **Number of references** | 76 (100) | 81 (100) | 74 (100) | 65 (100) | 68 (100) | 60 (100) |
| **Accurate references** | 22 (29)* | 31 (38)*** | 33 (45)*** | 9 (14)* | 7 (10)*** | 10 (17)*** |
| **All correct** | 10 (13) | 17 (21) | 19 (26) | 4 (6) | 1 (1) | 2 (3) |
| **Accurate but DOI number not given** | 12 (16) | 14 (17) | 14 (19) | 5 (8) | 6 (9) | 8 (13) |
| **Inaccurate references** | 54 (71) | 50 (62) | 41 (55) | 56 (86) | 61 (90) | 50 (83) |
| **Accurate but wrong DOI number** | 5 (7) | 3 (4) | 2 (3) | 0 (0) | 0 (0) | 1 (2) |
| **Partially accurate - missing some information (no false information)** | 24 (32) | 20 (25) | 23 (31) | 12 (18) | 11 (16) | 10 (17) |
| **Partially accurate but with some false information** | 20 (26) | 19 (23) | 12 (16) | 27 (42) | 43 (63) | 33 (55) |
| **False reference** | 5 (7) | 8 (10) | 4 (5) | 17 (26) | 7 (10) | 6 (10) |

\* $p < 0.05$ , \*\*\* $p < 0.001$

Several types of errors emerged when inaccuracies in the references were examined. First, there were errors only in the DOI number, which happened more often with ChatGPT. The change was often minor, such as just in the last digit, but it meant that a completely different paper was referenced. Next, some references were partially correct, with only some missing information, but more significant were those with additional false information (for example, with added incorrect authors). When examined more closely, it appears that these names can be found on the same page, and were often authors of other cited works. Sometimes, there were correct names but the order of the authors was wrong, and occasionally instead of the authors’ names the society to which they belong is given.

Finally, there were totally confected references, which occurred for 5–10% of those given by ChatGPT, and 10–26% for Gemini.

Table 3 shows the accuracy in terms of relevance to the paper that was used as the basis for the question. As the question directly asked for ‘guidelines’, one might expect that among the references suggested by the chatbot will be highly cited ones (i.e. those from Table 1). In fact, the percent of responses that contained these core references ranged from 20% to 48% for ChatGPT and from 20% to 24% for Gemini. The percent of core references that were found across all three sessions was 12% for ChatGPT and 4% for Gemini. Consistency analysis showed fair agreement for ChatGPT, and slight agreement for Gemini.

**Table 3.**
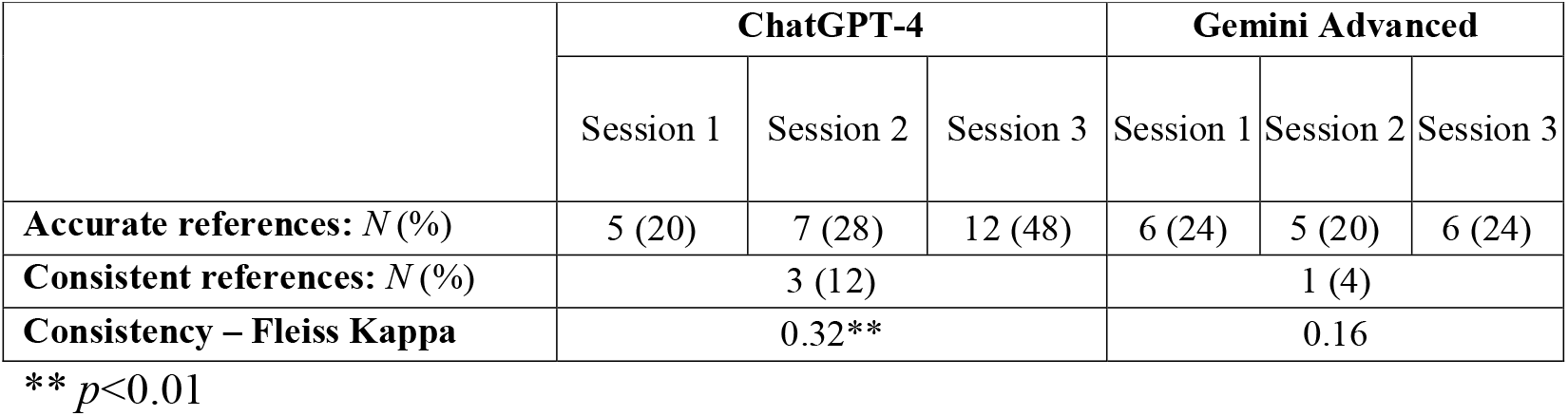
Accuracy related to the initial search question (i.e. whether any of the references found by the chatbot included the original paper from Table 1 on which the question was framed). The results for each session were compared (ChatGPT-4 vs Gemini Advanced) using Chi-squared tests, but there were no significant differences.

|  | ChatGPT-4 |  |  | Gemini Advanced |  |  |
| --- | --- | --- | --- | --- | --- | --- |
|  | Session 1 | Session 2 | Session 3 | Session 1 | Session 2 | Session 3 |
| **Accurate references: *N* (%)** | 5 (20) | 7 (28) | 12 (48) | 6 (24) | 5 (20) | 6 (24) |
| **Consistent references: *N* (%)** | 3 (12) |  |  | 1 (4) |  |  |
| **Consistency – Fleiss Kappa** | 0.32** |  |  | 0.16 |  |  |

\*\* $p < 0.01$

We also analyzed the percent of errors in each response (averaged across three sessions) in terms of the number of citations of each paper on which the question was based, and the results are shown in Fig. 2. For both chatbots there was a trend showing that a lower percentage of errors was associated with a higher number of citations. For ChatGPT the correlation was significant, but for Gemini it was not.

**Fig. 1.**
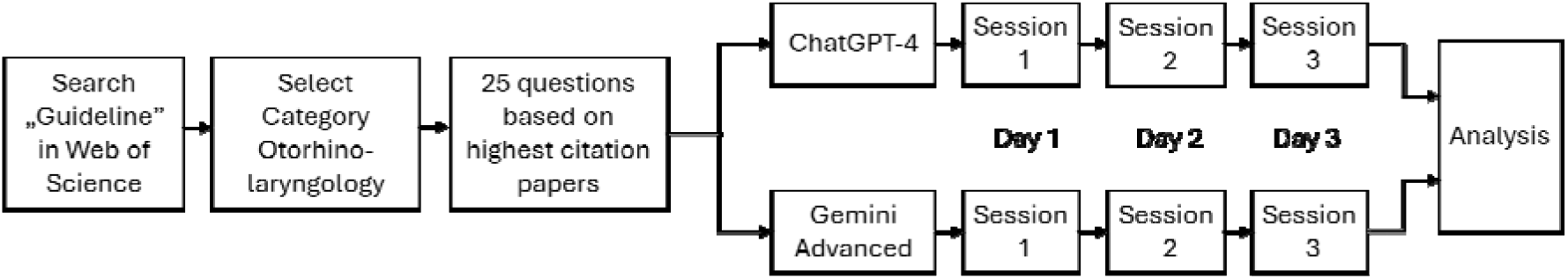
Diagram showing the protocol of the study.

**Fig. 2.**
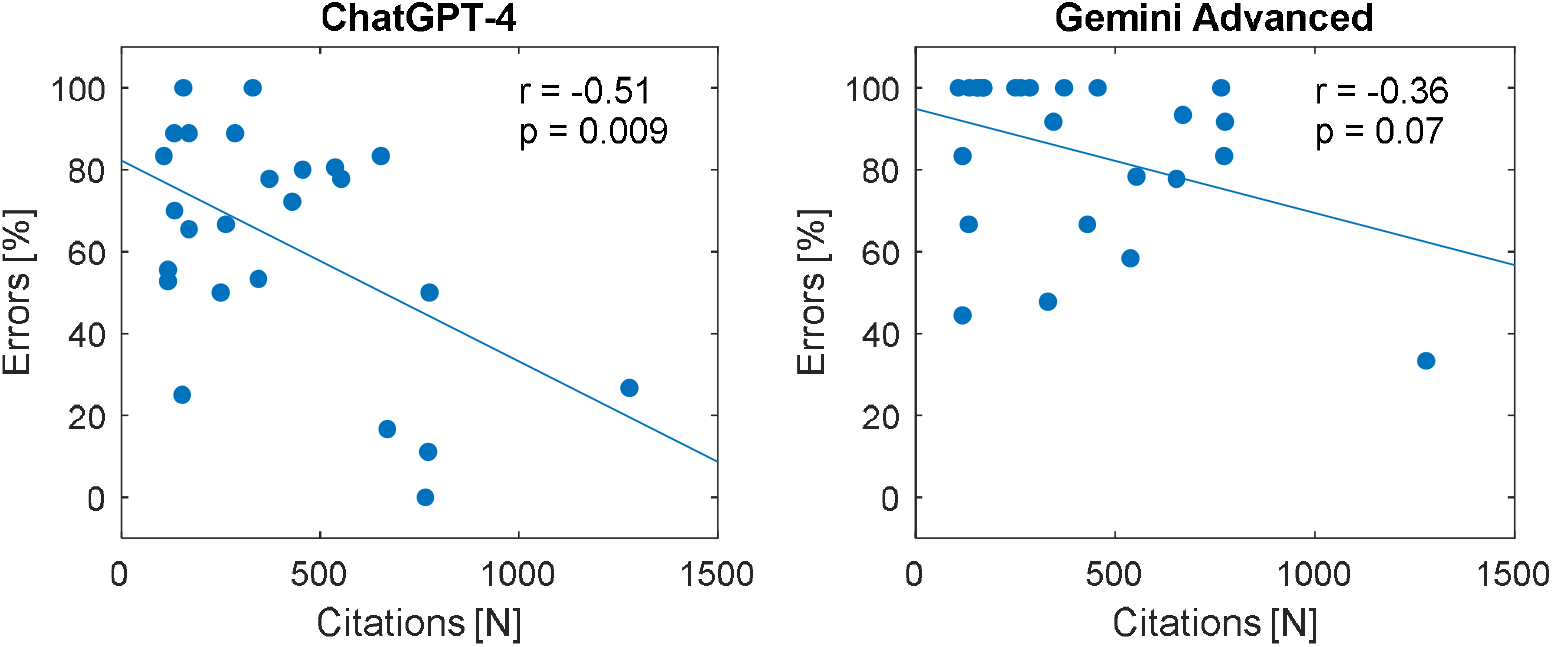
The percent of errors related to the number of citations for ChatGPT (left) and Gemini (right). *r* – correlation coefficient; *p* – level of significance.

## Discussion

Until now it was not known whether Gemini made up fake references in a similar way to what ChatGPT was known to do. However, as a starting point, it was known that the earlier version of Gemini, called Bard, did have that weakness [36, 37]. Gemini and Gemini Advanced are more sophisticated successors to Bard, and so it might be hoped that some progress has been made. Unfortunately, the present study shows that Gemini still generates false references.

In general, the present study shows that the accuracy of references provided by the best available models of ChatGPT and Gemini are still very poor. Previous studies have already shown that free versions have lesser capabilities and apparently perform worse [6, 7]. The results of the present study have shown that correct references are only given sporadically and that the overall performance is also made worse by low consistency. That raises the question: if chatbots are so poor when the information that is sought can be verified, how poor are they in other cases? Perhaps when their responses are being rated by experts the correct figure is in fact overestimated?

In the present study we not only classified responses as correct or incorrect but also checked what was correct and what was not. Common errors included: omissions of information, false author names, false DOI numbers, and completely fabricated references. Previous studies on references retrieved by chatbots have not mentioned any problem with DOI numbers [6, 7]. Our study has revealed the way in which chatbots make errors. The difference in a DOI number is often small, like changing the last digit, but the resulting error is actually serious because the mistaken DOI points to a different paper. The underlying reason might be because the DOIs are totally fabricated by the chatbot, or maybe the number was found on the same page containing titles of other papers, e.g. a table of contents.

An important result of the present study is that the percentage of errors correlated negatively with the number of citations. This reveals something that may seem obvious and expected, namely that chatbots perform better the more information they have. But what is not so obvious is that they apparently need some mechanism that stops them from falsifying information in areas where they are ill-trained. It is better for a user to receive the response “I don’t know” than to be misled.

This study further confirms that there is considerable variability in the results provided by chatbots [38]. The responses of both ChatGPT and Gemini varied across the three sessions. ChatGPT appears to have improved, a feature that has also been noted by some earlier studies, but it is not easy to confirm given the large variability [7, 39].

The poorer results than have been found in previous studies on otolaryngology references [6, 7] might be connected with several issues. The first is that the papers we used as the basis for our tests had fewer citations than in the study by Lechien [7]. Hence, there is less information in the training space used for chatbots and so more errors, as illustrated in Fig. 2.

## Conclusions

The present study shows that both ChatGPT and Gemini are unsuitable for retrieving references from the scientific literature, even though ChatGPT performs noticeably better than Gemini. This finding casts serious doubts on the correctness of the information provided by chatbots in general. However, we did find that the percentage of errors did decrease when there were larger numbers of citations. This probably relates to the fact that the more literature there is on a topic the more capable is the chatbot. Finally, our work indicates that while chatbots might perform well in broad domains of knowledge, they perform extremely poorly, and falsify information, in more specialized areas.

## Data Availability

All data produced in the present study are available as supplementary files.

