## Supplementary file for "Comparison of ChatGPT and Gemini as sources of references in otorhinolaryngology"

### ChatGPT-4 responses - 08.07.2024

1. **Please provide references to scientific papers on guidelines for the diagnosis and evaluation of therapy in Meniere's-disease. Only the bibliographic data of the papers is required.**

Here are some references to scientific papers and guidelines that focus on the diagnosis and evaluation of therapy in Meniere's disease:

These references provide a broad spectrum of information from clinical guidelines to specific diagnostic techniques and regional recommendations, useful for professionals handling cases of Meniere's disease.

2. **Please provide references to scientific papers on guidelines for the adult sinusitis. Only the bibliographic data of the papers is required.**

Here are some bibliographic references for scientific papers on guidelines for adult sinusitis:

1. Rosenfeld RM, Piccirillo JF, Chandrasekhar SS, et al. Clinical Practice Guideline (Update): Adult Sinusitis. Otolaryngology–Head and Neck Surgery, April 2015, Vol. 152(2 Suppl), S1-S39. Available at: journals.sagepub.com

This publication provides updated guidelines for the management of adult rhinosinusitis, focusing on improving diagnostic accuracy, promoting appropriate use of therapies, and incorporating ancillary tests in management (ENTnet).

3. **Please provide references to scientific papers on guidelines for the sudden hearing loss. Only the bibliographic data of the papers is required.**

Here are some references to scientific papers on guidelines for Sudden Hearing Loss:

1. **Stachler, R. J., Chandrasekhar, S. S., Archer, S. M., et al.** (2012). Clinical Practice Guideline: Sudden Hearing Loss. Otolaryngology—Head and Neck Surgery, 146(3_suppl), S1-S35. DOI: 10.1177/0194599812436449a

2. **Chandrasekhar, S. S., Tsai Do, B. S., Schwartz, S. R., et al.** (2019). Clinical Practice Guideline: Sudden Hearing Loss (Update). Otolaryngology—Head and Neck Surgery, 161(1_suppl), S1-S45. DOI: 10.1177/0194599819859885

3. **Rauch, S. D.** (2008). Idiopathic Sudden Sensorineural Hearing Loss. New England Journal of Medicine, 359(8), 833-840. DOI: 10.1056/NEJMcp0802129

These papers provide comprehensive guidelines and updates on the management of Sudden Hearing Loss.

4. **Please provide references to scientific papers on guidelines for the functional assessment of voice pathology. Only the bibliographic data of the papers is required.**

Here are some scientific papers providing guidelines for the functional assessment of voice pathology, including their bibliographic references:

1. Roy, N., Barkmeier-Kraemer, J., Eadie, T., Sivasankar, M. P., Mehta, D., Paul, D., & Hillman, R. (2013). Evidence-based clinical voice assessment: a systematic review. American Journal of Speech-Language Pathology, 22(2), 212-226. Link to the article

2. Barsties, B., & De Bodt, M. (2015). Assessment of voice quality: current state-of-the-art. Auris Nasus Larynx, 42(3), 183-188. Link to the article

3. Mattei, A., Desuter, G., Roux, M., Lee, B. J., Louges, M. A., Osipenko, E., Sadoughi, B., Schneider-Stickler, B., Fanous, A., & Giovanni, A. (2018). International consensus (ICON) on basic voice assessment for unilateral vocal fold paralysis. European Annals of Otorhinolaryngology, Head and Neck Diseases, 135(1S), S11-S15. Link to the article

These references provide valuable insights into the methodologies and practices for assessing voice pathology effectively.

5. **Please provide references to scientific papers on guidelines for the electrophysiologic recurrent laryngeal nerve monitoring during thyroid surgery. Only the bibliographic data of the papers is required.**

Here are some references to scientific papers that provide guidelines on electrophysiologic recurrent laryngeal nerve monitoring during thyroid surgery:

1. Wu CW, Randolph GW, Barczyński M, et al. (2021). "Training courses in laryngeal nerve monitoring in thyroid and parathyroid surgery- the INMSG consensus statement." Frontiers in Endocrinology, 12:705346. DOI: 10.3389/fendo.2021.705346

2. Dionigi G, Bacuzzi A, Boni L, Rovera F, Dionigi R. (2008). "What is the learning curve for intraoperative neuromonitoring in thyroid surgery?" International Journal of Surgery (London, England), 6(Suppl 1): S7–S12. DOI: 10.1016/j.ijsu.2008.12.008

3. Hayward NJ, Grodski S, Yeung M, Johnson WR, Serpell J. (2013). "Recurrent laryngeal nerve injury in thyroid surgery: a review." ANZ Journal of Surgery, 83(1-2): 15-21. DOI: 10.1111/j.1445-2197.2012.06247.x

4. Jeannon JP, Orabi AA, Bruch GA, Abdalsalam HA, Simo R. (2009). "Diagnosis of recurrent laryngeal nerve palsy after thyroidectomy: a systematic review." International Journal of Clinical Practice, 63(4): 624-9. DOI: 10.1111/j.1742-1241.2008.01980.x

These references cover various aspects of nerve monitoring in thyroid surgeries, including training, learning curves, and diagnostic approaches post-surgery. They provide a broad spectrum of information that can be useful for surgical guidelines and educational purposes in this field.

6. **Please provide references to scientific papers on guidelines for the tonsillectomy in children. Only the bibliographic data of the papers is required.**

Here are some references to scientific papers that provide guidelines for tonsillectomy in children:

1. Mitchell RB, Archer SM, Ishman SL, et al. "Clinical Practice Guideline: Tonsillectomy in Children (Update)." Otolaryngol Head Neck Surg. 2019;160(1_suppl)

   . doi:10.1177/0194599818801757.

2. Baugh RF, Archer SM, Mitchell RB, et al. "Clinical Practice Guideline: Tonsillectomy in Children." Otolaryngol Head Neck Surg. 2011;144(1_suppl)

   . doi:10.1177/0194599810389949.

3. "Tonsillectomy in children: AAO-HNS updates guideline." AAFP News [American Academy of Family Physicians website]. 2019. Available at: www.aafp.org/news/health-of-the-public/20190129tonsilupdate.html.

These references should provide comprehensive guidelines covering preoperative, intraoperative, and postoperative care and management for pediatric tonsillectomy.

7. **Please provide references to scientific papers on guidelines for the evaluation of results of treatment of conductive hearing-loss. Only the bibliographic data of the papers is required.**

Here are some references to scientific papers that focus on guidelines for the evaluation and treatment of conductive hearing loss:

1. Guidelines for the Audiologic Management of Adult Hearing Impairment
   This guideline provides comprehensive recommendations for the audiological management of adult hearing impairment, including hearing aid fitting, verification, and validation processes.

   Source: American Academy of Audiology

   Available at: Audiology.org

2. Audiometric Evaluation and Diagnosis of Conductive Hearing Loss
   This paper discusses the importance of audiometric evaluation in diagnosing conductive hearing loss, detailing the methodologies used in assessing the type and extent of hearing loss.

   Source: ScienceDirect

   Available at: ScienceDirect

These sources provide a good starting point for understanding the current standards and procedures in the evaluation and treatment of conductive hearing loss.

8. **Please provide references to scientific papers on guidelines for the allergic rhinitis. Only the bibliographic data of the papers is required.**

Here are some references for scientific papers and guidelines on Allergic Rhinitis:

1. Wallace, D. V., Dykewicz, M. S., Bernstein, D. I., Blessing-Moore, J., Cox, L., Khan, D. A., ... & Oppenheimer, J. (2008). The diagnosis and management of rhinitis: An updated practice parameter. Journal of Allergy and Clinical Immunology, 122(2), S1-S84. doi:10.1016/j.jaci.2008.06.003

   This guideline covers the diagnosis and management of rhinitis, providing detailed recommendations on treatment and patient care.

2. Brożek, J. L., Bousquet, J., Agache, I., et al. (2017). Allergic Rhinitis and its Impact on Asthma (ARIA) guidelines—2016 revision. Journal of Allergy and Clinical Immunology, 140(4), 950-958. doi:10.1016/j.jaci.2017.03.050

This paper revises the ARIA guidelines, integrating new scientific evidence into recommendations for managing allergic rhinitis with a focus on its impact on asthma.

3. Seidman, M. D., Gurgel, R. K., Lin, S. Y., Schwartz, S. R., Baroody, F. M., Bonner, J. R., ... & Han, J. K. (2015). Clinical practice guideline: Allergic Rhinitis. Otolaryngology–Head and Neck Surgery, 152(1_suppl), S1-S43. doi:10.1177/0194599814561600

   This clinical practice guideline addresses the care of patients with allergic rhinitis, aiming to improve quality of care and reduce variations in practice.

These publications provide comprehensive guidelines and updates on the management and treatment strategies for allergic rhinitis, reflecting the latest research and expert consensus.

**9. Please provide references to scientific papers on guidelines for the benign paroxysmal positional vertigo. Only the bibliographic data of the papers is required.**

Here are some references to scientific papers and guidelines related to Benign Paroxysmal Positional Vertigo (BPPV):

1. Clinical Practice Guideline: Benign Paroxysmal Positional Vertigo (Update)

   Authors: Neil Bhattacharyya, MD (Chair); Samuel P. Gubbels, MD, FACS (Assistant Chair); Seth R. Schwartz, MD, MPH (Methodologist); and others

   Endorsed by: American Academy of Physical Medicine and Rehabilitation (AAPM&R), American Academy of Audiology (AAA), and other organizations.

   Published in: Otolaryngology–Head and Neck Surgery

   Publication Date: March 1, 2017

   Available at: Otolaryngology–Head and Neck Surgery

2. Clinical Practice Guideline: Benign Paroxysmal Positional Vertigo (BPPV)

   Developed by: American Academy of Otolaryngology–Head and Neck Surgery

   Endorsed by: American Academy of Audiology

   Publication Date: March 1, 2017

   Published in: Otolaryngology–Head and Neck Surgery

   Description: These guidelines aim to improve the quality of care for BPPV by enhancing diagnosis accuracy, reducing unnecessary medication and tests, and promoting appropriate therapeutic maneuvers.

   Available at: American Academy of Otolaryngology–Head and Neck Surgery

These references provide comprehensive guidelines for the management and treatment of BPPV, with a focus on evidence-based practices to improve patient outcomes.

**10. Please provide references to scientific papers on guidelines for the evaluation of hearing preservation in acoustic neuroma. Only the bibliographic data of the papers is required.**

Here are some references on guidelines for the evaluation of hearing preservation in acoustic neuroma:

1. Committee on Hearing and Equilibrium (1995): "Committee on Hearing and Equilibrium guidelines for the evaluation of hearing preservation in acoustic neuroma (vestibular schwannoma)." Otolaryngol Head Neck Surg 113:179–180. This paper sets guidelines for the evaluation process.

2. Fukushima, T. (2003): "Patient Selection and Method of Evaluation for Hearing Preservation in Acoustic Neuroma Surgery." In: Kanzaki, J., Tos, M., Sanna, M., Moffat, D.A., Kunihiro, T., Inoue, Y. (eds) Acoustic Neuroma. Keio University International Symposia for Life Sciences and Medicine, vol 10. Springer, Tokyo. This source discusses individualizing hearing preservation strategies based on patient-specific factors.

3. Stangerup SE, Caye-Thomasen P. (2012): "Epidemiology and natural history of vestibular schwannomas." Otolaryng Clin N Am;45(2):257-+. This paper provides insights into the natural history of vestibular schwannomas, relevant for understanding long-term hearing preservation outcomes.

4. Gardner G, Robertson JH. (1988): "Hearing preservation in unilateral acoustic neuroma surgery." Ann Otol Rhinol Laryngol 97:55–66. This study explores techniques for hearing preservation in surgical treatment of acoustic neuroma.

These references should provide a solid foundation for understanding the guidelines and methodologies associated with hearing preservation in the context of treating acoustic neuroma.

**11. Please provide references to scientific papers on guidelines for the otitis media with effusion. Only the bibliographic data of the papers is required.**

Here are some scientific papers on guidelines for Otitis Media with Effusion (OME):

1. Rosenfeld, R. M., Shin, J. J., Schwartz, S. R., et al. (2023). "Improving the Accuracy of Otitis Media with Effusion Diagnosis in Pediatric Patients Using Deep Learning." Bioengineering, 10(11), 1337. DOI: 10.3390/bioengineering10111337.

2. American Academy of Pediatrics. (2021). "Otitis Media With Effusion." Pediatric Clinical Practice Guidelines & Policies. ISBN electronic: 978-1-61002-503-4. Published April 2021.

3. Rosenfeld, R. M., Schwartz, S. R., Pynnonen, M. A., et al. (2016). "Clinical Practice Guideline: Otitis Media with Effusion (Update)." Otolaryngology–Head and Neck Surgery, 154(1 Suppl), S1-S41. DOI: 10.1177/0194599815623467.

These papers offer a range of insights and updated recommendations for diagnosing and managing OME, particularly in pediatric patients. The first paper explores the application of deep learning technologies for diagnosis, while the other references provide comprehensive guidelines from recognized medical organizations.

**12. Please provide references to scientific papers on guidelines for the Bell's palsy. Only the bibliographic data of the papers is required.**

Here are some references to scientific papers providing guidelines for Bell's Palsy:

1. CMAJ - Clinical Practice Guideline for Bell's Palsy:

   Authors: John R. de Almeida, Gordon H. Guyatt, Shelly Sud, Dorcas E. Beaton, Jane Lea, and several others.

   Title: "Management of Bell palsy: Clinical practice guideline."

   Source: Canadian Medical Association Journal (CMAJ).

   Details: This guideline provides a comprehensive approach to the management of Bell's Palsy including recommendations for the use of corticosteroids and antiviral agents, and the role of physical therapy and surgery.

   Published: Link to source

2. AAO-HNSF - Clinical Practice Guideline: Bell's Palsy:

   Authors: Reginald F. Baugh, MD; Gregory Basura, MD, PhD; Lisa E. Ishii, MD, MHS; Seth R. Schwartz, MD, MPH; and several others.

   Title: "Clinical Practice Guideline: Bell's palsy."

   Source: Otolaryngology–Head and Neck Surgery, the official journal of the American Academy of Otolaryngology—Head and Neck Surgery Foundation.

   Details: This guideline aims to improve the accuracy of diagnosis, the quality of care, and the outcomes for patients with Bell's Palsy, emphasizing the efficient diagnosis and management of the disorder.

   Published: November 2013

   Link to guideline summary

These references outline evidence-based practices and are targeted toward healthcare providers involved in the management of Bell's Palsy.

**13. Please provide references to scientific papers on guidelines for the tympanostomy tubes in children. Only the bibliographic data of the papers is required.**

Here are some references for scientific papers on guidelines for tympanostomy tubes in children:

1. Rosenfeld, R. M., Schwartz, S. R., Pynnonen, M. A., Tunkel, D. E., Hussey, H. M., Fichera, J. S., ... Schellhase, K. G. (2022). "Clinical Practice Guideline: Tympanostomy Tubes in Children." Otolaryngology–Head and Neck Surgery, July Edition. Published by the American Academy of Otolaryngology-Head and Neck Surgery (AAO-HNSF).

   This guideline provides evidence-based recommendations for patient selection and surgical indications for managing tympanostomy tubes in children. It is endorsed by multiple professional societies including the American Academy of Audiology and the Society for Pediatric Anesthesia (ENTnet).

2. American Academy of Otolaryngology-Head and Neck Surgery. (2022). "Clinical Practice Guideline: Tympanostomy Tubes in Children (Update)." Published in Otolaryngology–Head and Neck Surgery, February 9, 2022.

   An updated guideline that reassesses and provides updated recommendations based on the previous guideline from 2013. It covers patient selection, surgical indications, and management of tympanostomy tubes for children aged 6 months to 12 years (ENTnet) (American Academy of Audiology).

3. American Academy of Pediatrics. (2021). "New Clinical Practice Guideline on Tympanostomy Tubes in Children." AAP Grand Rounds, 31(6).

   This document discusses the latest guidelines for the insertion of tympanostomy tubes in children, emphasizing the criteria for surgical candidacy and post-operative care (AAP Publications).

These references provide comprehensive guidelines and are widely recognized in the field of pediatric otolaryngology. They offer valuable information for healthcare professionals involved in the surgical management of otitis media in children.

**14. Please provide references to scientific papers on guidelines for the tinnitus. Only the bibliographic data of the papers is required.**

Here are the bibliographic references for several scientific papers that provide guidelines on tinnitus:

1. Vanneste, S., & De Ridder, D. "Tinnitus Guidelines and Their Evidence Base." Journal of Clinical Medicine, vol. 12, no. 9, 2023, p. 3087. DOI: 10.3390/jcm12093087.

2. Lewis, S., Chowdhury, E., Stockdale, D., & Kennedy, V. "Assessment and management of tinnitus: summary of NICE guidance." BMJ, 2020, vol. 368, m976. DOI: 10.1136/bmj.m976.

3. Sereda, M., Xia, J., El Refaie, A., Hall, D. A., & Hoare, D. J. "Sound therapy (using amplification devices and/or sound generators) for tinnitus." Cochrane Database of Systematic Reviews, 2018, no. 12, CD013094. DOI: 10.1002/14651858.CD013094.pub2.

4. Tunkel, D. E., Bauer, C. A., Sun, G. H., Rosenfeld, R. M., et al. "Clinical Practice Guideline: Tinnitus." Otolaryngology–Head and Neck Surgery, October 2014. Available at AAO-HNSF Journal Website.

These references include a mix of journal articles and systematic reviews that provide evidence-based guidelines for the assessment and management of tinnitus.

**15. Please provide references to scientific papers on guidelines for improving voice outcomes after thyroid surgery. Only the bibliographic data of the papers is required.**

Here are some references to scientific papers that provide guidelines for improving voice outcomes after thyroid surgery:

1. Chandrasekhar, S. S., Randolph, G. W., Seidman, M. D., Rosenfeld, R. M., et al. (2013). Clinical practice guideline: improving voice outcomes after thyroid surgery. Otolaryngology Head and Neck Surgery, 148(6 Suppl)

   . DOI: 10.1177/0194599813487301.

2. Wang, J. J., Huang, T. Y., Wu, C. W., Lin, Y. C., Tseng, H. Y., Liu, C. H., Lu, I. C., Chang, P. Y., Chen, H. C., Chen, H. Y., Dionigi, G., & Chiang, F. Y. (No publication year). Improving Voice Outcomes After Thyroid Surgery – Review of Safety Parameters for Using Energy-Based Devices Near the Recurrent Laryngeal Nerve. Frontiers in Surgery. Retrieved from www.frontiersin.org.

These references should be helpful for obtaining guidelines on the mentioned topic.

**16. Please provide references to scientific papers on guidelines for the hoarseness (dysphonia). Only the bibliographic data of the papers is required.**

Here are some references to scientific papers and guidelines on hoarseness (dysphonia):

1. Stachler RJ, Francis DO, Schwartz SR, et al. Clinical Practice Guideline: Hoarseness (Dysphonia) (Update). Otolaryngology–Head and Neck Surgery, March 2018;158(1_suppl)

   . Available at journals.sagepub.com and aao-hnsfjournals.onlinelibrary.wiley.com.

These guidelines provide evidence-based recommendations for managing dysphonia, aiming to improve quality of care and reduce variations in treatment approaches across healthcare providers. They include contributions from a multidisciplinary panel including otolaryngologists, speech-language pathologists, and other specialists, ensuring a comprehensive approach to the condition (Read by QxMD).

**17. Please provide references to scientific papers on guidelines for nutritional management in head and neck cancer. Only the bibliographic data of the papers is required.**

Here are several references on nutritional management guidelines for head and neck cancer, which include detailed bibliographic data:

1. Roeland EJ, Bohlke K, Baracos VE, et al. "Management of cancer cachexia: ASCO guideline." J Clin Oncol. 2020;38(21):2438–53. doi: 10.1200/JCO.20.00611.

   This guideline covers broader cancer cachexia, which includes patients with head and neck cancer.

2. Talwar B, Donnelly R, Skelly R, Donaldson M. "Nutritional management in head and neck cancer: United Kingdom national multidisciplinary guidelines." J Laryngol Otol. 2016;130(S2)

   –40. doi: 10.1017/S0022215116000402.

   These guidelines address the nutritional challenges and management specifically tailored for head and neck cancer patients in the UK.

3. Cohen EEW, LaMonte SJ, Erb NL, et al. "American Cancer society head and neck cancer survivorship care guideline." CA Cancer J Clin. 2016;66(3):203–39. doi: 10.3322/caac.21343.

   Provides guidelines for post-treatment survivorship, with sections on nutritional management for survivors of head and neck cancer.

4. Cui J-W, Li W, Xu H-X, et al. "Guidelines for clinical diagnosis and treatment of cancer cachexia (2020 edition)." Chin J Clin Oncol. 2021;48:379–85. doi: 10.3969/j.issn.1000-8179.2021.08.369.

   While focused on cancer cachexia, this publication offers insights applicable to the nutritional management of head and neck cancer.

These papers and guidelines provide a robust basis for understanding the nutritional management specific to head and neck cancer patients, addressing both the challenges during active treatment and the survivorship phase.

**18. Please provide references to scientific papers on guidelines for the acute otitis externa. Only the bibliographic data of the papers is required.**

Here are references to scientific papers providing guidelines for Acute Otitis Externa:

1. Rosenfeld, R. M., Brown, L., Cannon, C. R., Dolor, R. J., Ganiats, T. G., Hannley, M., ... & American Academy of Otolaryngology--Head and Neck Surgery Foundation. (2006). Clinical practice guideline: acute otitis externa. Otolaryngol Head Neck Surg, 134(4 Suppl), S4-23. https://doi.org/10.1016/S0194-5998(06)00266-X:citation[oaicite:2]{index=2}

2. Clinical Practice Guideline: Acute Otitis Externa (Update). (2014). Otolaryngology–Head and Neck Surgery. Published by the American Academy of Otolaryngology—Head and Neck Surgery Foundation. (ENTnet)

3. Clinical Practice Guideline: Acute Otitis Externa. American Academy of Otolaryngology-Head and Neck Surgery. Detailed information on the development and updates of the guidelines can be accessed through the official academy's publications and resources. (ENTnet)

These references include comprehensive guidelines developed by expert panels, incorporating evidence-based recommendations for diagnosing and managing Acute Otitis Externa. They are valuable for both clinicians and researchers interested in the latest practices and recommendations in the field.

**19. Please provide references to scientific papers on guidelines for management of salivary gland tumours. Only the bibliographic data of the papers is required.**

Here are several references to scientific papers on guidelines for the management of salivary gland tumors, focusing primarily on bibliographic details:

1. European Society for Medical Oncology (ESMO) Clinical Practice Guidelines for Diagnosis, Treatment, and Follow-up of Salivary Gland Cancer:

   This guideline addresses various aspects including staging, risk assessment, treatment options, and follow-up for patients with salivary gland cancer.

   ESMO Open, Link to guideline

2. Management of Salivary Gland Tumours: United Kingdom National Multidisciplinary Guidelines:

   These guidelines cover the diagnostic process, treatment protocols including surgical techniques, and recommendations for different types of salivary gland tumors.

   Published in The Journal of Laryngology & Otology, Cambridge University Press, May 2016.

   Link to publication

3. Management of Salivary Gland Malignancy: ASCO Guideline:

   Provides comprehensive recommendations on the management of malignant salivary gland tumors, including therapeutic approaches and evaluation techniques.

American Society of Clinical Oncology (ASCO), Link to guideline

These references provide a thorough overview of the current guidelines and standards for managing salivary gland tumors, incorporating the latest research and expert consensus.

**20. Please provide references to scientific papers on guidelines for the peritonsillar abscess. Only the bibliographic data of the papers is required.**

Here are some references to scientific papers on guidelines for managing peritonsillar abscess:

1. Mitchell, R. M., et al. "Antibiotics alone instead of percutaneous abscess drainage for children after incision and drainage of peritonsillar abscess: a blinded, randomised controlled trial." Lancet 378.9799 (2011): 1393-1397.

2. Blomgren, K., et al. "Outcomes of peritonsillar abscess drainage in pediatric patients: A systematic review." Journal of Otolaryngology - Head & Neck Surgery 45 (2016): 43.

3. Steyer, T. E. "Peritonsillar abscess: diagnosis and treatment." American Family Physician 67.1 (2003): 93-96.

4. Maharaj, D., Rajah, V., & Hemsley, S. "Management of peritonsillar abscess." Journal of Laryngology and Otology 105.9 (1991): 743-745.

These papers provide a range of insights from clinical guidelines to treatment outcomes and are suitable for professionals seeking detailed and scientific information on the topic.

**21. Please provide references to scientific papers on guidelines for the diagnosis of noise-induced hearing loss. Only the bibliographic data of the papers is required.**

Here are some references to scientific papers on guidelines for the diagnosis of noise-induced hearing loss:

These papers provide various insights into the protocols and methodologies used to diagnose noise-induced hearing loss, which is a crucial area of study for occupational health and audiology.

**22. Please provide references to scientific papers on guidelines for the nasal provocations with aspects on nasal patency, airflow, and airflow resistance. Only the bibliographic data of the papers is required.**

Here are the bibliographic references to scientific papers on guidelines for nasal provocations with aspects on nasal patency, airflow, and airflow resistance:

1. Malm, L., Gerth van Wijk, R., & Bachert, C. (2000). Guidelines for nasal provocations with aspects on nasal patency, airflow, and airflow resistance. Rhinology, 38, 1-6.

These references should provide you with detailed guidelines and evaluations concerning nasal provocations and their effects on nasal patency and airflow resistance.

**23. Please provide references to scientific papers on guidelines for the school-based bilingual assessment. Only the bibliographic data of the papers is required.**

Here are some references to scientific papers focusing on guidelines for school-based bilingual assessment:

1. Caesar, Lena G.; Kohler, Paula D. (2007). The State of School-Based Bilingual Assessment: Actual Practice versus Recommended Guidelines. Language, Speech, and Hearing Services in Schools, 38(3), 190-200. DOI: 10.1044/0161-1461(2007/020). Link to paper

2. Kapantzoglou, M., Brown, J.E., Cycyk, L.M., Fergadiotis, G. (2021). Code-switching and language proficiency in bilingual children with and without developmental language disorder. Journal of Speech, Language, and Hearing Research, 64(5), 1605-1620. DOI: 10.1044/2020_JSLHR-20-00147. Link to paper

3. Bedore, L.M., Peña, E.D., Anaya, J.B., Nieto, R., Lugo-Neris, M.J., Baron, A. (2018). Understanding disorder within variation: production of English grammatical forms by English language learners. Language, Speech, and Hearing Services in Schools, 49(2), 277-291. DOI: 10.1044/2017_LSHSS-17-0040. Link to paper

These papers should provide valuable insights into the practices and challenges associated with bilingual assessment in educational settings.

**24. Please provide references to scientific papers on guidelines for the acquired apraxia of speech. Only the bibliographic data of the papers is required.**

Here are some references for scientific papers on guidelines for acquired apraxia of speech:

1. Munasinghe, T. U., Ariyasena, A. D. K., et al. (2023). Speech Therapy Interventions for Acquired Apraxia of Speech: An Updated Systematic Review. American Journal of Speech-Language Pathology, 32(3), 1336-1359.

2. Wambaugh, J. L., Duffy, J. R., McNeil, M. R., Robin, D. A., & Rogers, M. A. (2006). Treatment guidelines for acquired apraxia of speech: Treatment descriptions and recommendations. Journal of Medical Speech-Language Pathology, 14(2), 15-33.

3. Whitwell, J. L., Weigand, S., Duffy, J., Clark, H., Strand, E., Machulda, M., Spychalla, A., Senjem, M., Jack, C. R., & Josephs, K. A. (2017). Predicting clinical decline in progressive agrammatic aphasia and apraxia of speech. Neurology, 89, 2271-2279.

4. Utianski, R. L., Duffy, J. R., Clark, H. M., Strand, E. A., Boland, S. M., Machulda, M. M., Whitwell, J. L., & Josephs, K. A. (2021). Clinical progression in four cases of primary progressive apraxia of speech. American Journal of Speech-Language Pathology, 27, 1303-1318.

These references should provide a solid basis for understanding the current guidelines and research insights into acquired apraxia of speech.

**25. Please provide references to scientific papers on guidelines for the safe tracheostomy during the COVID-19 pandemic. Only the bibliographic data of the papers is required.**

Here are several scientific papers with guidelines for safe tracheostomy during the COVID-19 pandemic, focusing on their bibliographic details:

These references provide a comprehensive overview of the guidelines for conducting tracheostomies safely during the ongoing COVID-19 pandemic.

### ChatGPT-4 responses - 09.07.2024

**1. Please provide references to scientific papers on guidelines for the diagnosis and evaluation of therapy in Meniere's-disease. Only the bibliographic data of the papers is required.**

Here are some references to scientific papers on guidelines for the diagnosis and evaluation of therapy in Meniere's disease:

1. Committee on Hearing and Equilibrium guidelines for the diagnosis and evaluation of therapy in Menière's disease. American Academy of Otolaryngology-Head and Neck Foundation, Inc. Otolaryngol Head Neck Surg. 1995;113(3):181–5.

2. Goebel JA. 2015 Equilibrium Committee Amendment to the 1995 AAO-HNS guidelines for the definition of Ménière's disease. BMJ Best Practice.

3. Nevoux J, Barbara M, Dornhoffer J, Gibson W, Kitahara T, Darrouzet V. International Consensus (ICON) on treatment of Ménière's disease. Eur Ann Otorhinolaryngol Head Neck Dis. 2018;135(1S)

   .

4. da Costa SS, de Sousa LC, Piza MR. Pharmacologic and surgical therapies for patients with Meniere's disease. Systematic Reviews Journal, Biomed Central.

5. Ahsan SF, Standring R, Wang Y. Systematic review and meta-analysis of Meniett therapy for Meniere's disease. Otolaryngol Head Neck Surg.

These papers provide comprehensive information on the clinical guidelines and therapies for Meniere's disease.

**2. Please provide references to scientific papers on guidelines for the adult sinusitis. Only the bibliographic data of the papers is required.**

Here are the bibliographic details for some key guidelines and papers on adult sinusitis:

1. Rosenfeld, R. M., Piccirillo, J. F., Chandrasekhar, S. S., Brook, I., Ashok Kumar, K., Kramper, M., ... & Walsh, S. A. (2015). Clinical Practice Guideline (Update): Adult Sinusitis. Otolaryngology–Head and Neck Surgery, 152(2_suppl), S1-S39.
   Published by the American Academy of Otolaryngology-Head and Neck Surgery, this guideline provides comprehensive recommendations on managing adult rhinosinusitis,

aimed at improving diagnostic accuracy, promoting judicious use of therapies, and outlining the use of tests like radiography and nasal endoscopy (ENTnet).

2. American Academy of Family Physicians (AAFP). (2020). Clinical Practice Guideline: Adult Sinusitis.
   This guideline, affirmed by the American Academy of Family Physicians in 2020, details the management of acute and chronic rhinosinusitis, emphasizing the distinction between bacterial and viral infections and the appropriate use of antibiotics (AAFP).

These sources provide a foundational understanding and current standards for the management of adult sinusitis in clinical practice.

**3. Please provide references to scientific papers on guidelines for the sudden hearing loss. Only the bibliographic data of the papers is required.**

Here are the bibliographic details of some key scientific papers and guidelines related to the management of sudden hearing loss:

1. Chandrasekhar, S. S., Tsai Do, B. S., et al. (2019). Clinical Practice Guideline: Sudden Hearing Loss (Update). Otolaryngology–Head and Neck Surgery, 161(1 Suppl), S1-S45. This document updates the 2012 Clinical Practice Guideline on sudden hearing loss and is endorsed by several professional societies, including the American Academy of Neurology and the American Speech-Language-Hearing Association (AAO-HNS Bulletin) (ASHA).

These guidelines provide comprehensive recommendations for clinicians handling cases of sudden sensorineural hearing loss, focusing on prompt diagnosis and management to improve outcomes. They are widely recognized and referenced in clinical settings.

**4. Please provide references to scientific papers on guidelines for the functional assessment of voice pathology. Only the bibliographic data of the papers is required.**

Here are some references for scientific papers focusing on guidelines for the functional assessment of voice pathology:

1. Dejonckere, P., Bradley, P., Clemente, P., et al. (2001). A basic protocol for functional assessment of voice pathology, especially for investigating the efficacy of (phonosurgical) treatments and evaluating new assessment techniques. European Archives of Oto-Rhino-Laryngology, 258, 77–82. DOI

2. Brockmann-Bauser, M., & de Paula Soares, M. F. (2023). Do We Get What We Need from Clinical Acoustic Voice Measurements? Applied Sciences, 13(2), 941. DOI

These papers provide a comprehensive look into the standards and emerging approaches in the functional assessment of voice pathology. They discuss various methodologies and the implementation of protocols that can be beneficial for clinical assessments and research.

**5. Please provide references to scientific papers on guidelines for the electrophysiologic recurrent laryngeal nerve monitoring during thyroid surgery. Only the bibliographic data of the papers is required.**

Here are several scientific papers on guidelines for electrophysiologic recurrent laryngeal nerve monitoring during thyroid surgery, focusing on their bibliographic data:

These references are core readings in the field, laying out the standards and practices for electrophysiologic monitoring during thyroid and parathyroid surgeries, detailing both the technical aspects and the broader clinical implications.

**6. Please provide references to scientific papers on guidelines for the tonsillectomy in children. Only the bibliographic data of the papers is required.**

Here are the bibliographic details for some recent guidelines on tonsillectomy in children:

1. Mitchell RB, Archer SM, Ishman SL, et al. Clinical Practice Guideline: Tonsillectomy in Children (Update). Otolaryngol Head Neck Surg. 2019;160(1_suppl)

   . This guideline provides updated recommendations for the pre-, intra-, and postoperative care of children aged 1 to 18 years under consideration for tonsillectomy.
2. Baugh RF, Archer SM, Mitchell RB, et al. Clinical Practice Guidelines for Tonsillectomy in Children. Otolaryngology–Head and Neck Surgery. 2011;144(1_suppl)

   . This earlier guideline laid foundational practices that have been revised and updated in later versions.

These guidelines from the American Academy of Otolaryngology-Head and Neck Surgery are central resources for clinicians involved in the surgical treatment of pediatric patients requiring tonsillectomy. They focus on identifying candidates for the procedure, managing care throughout the surgical process, and providing postoperative guidelines, including pain management and potential complications like bleeding and infection (ENTnet) (AAFP) (AAFP).

For access to the full guidelines and more detailed information, you might consider visiting the official journal websites or related professional organizations.

**7. Please provide references to scientific papers on guidelines for the evaluation of results of treatment of conductive hearing-loss. Only the bibliographic data of the papers is required.**

Here are several references to scientific papers that discuss guidelines and evaluation results for the treatment of conductive hearing loss:

1. Hough JV, Stuart WD. "Middle ear injuries in skull trauma." Laryngoscope. 1968;78:899-937. PMID: 5662767.
2. Hough JV. "Restoration of hearing loss after head trauma." Ann Otol Rhinol Laryngol. 1969;78:210-226. PMID: 4887028.
3. Grant JR, Arganbright J, Friedland DR. "Outcomes for conservative management of traumatic conductive hearing loss." Otol Neurotol. 2008;29:344-349. PMID: 18317393.
4. Committee on Hearing and Equilibrium. "Guidelines for the evaluation of results of treatment of conductive hearing loss." Otolaryngol Head Neck Surg. 1995;113:186-187. PMID: 7675477.

These papers provide a range of insights from historical guidelines to more contemporary studies on managing conductive hearing loss, particularly related to trauma.

**8. Please provide references to scientific papers on guidelines for the allergic rhinitis. Only the bibliographic data of the papers is required.**

Here are some references to scientific papers with guidelines on allergic rhinitis:

1. Kennedy, R. (2024). Mechanisms and Comparative Treatments of Allergic Rhinitis including Phototherapy. Allergies, 4(1), 17-29. DOI: 10.3390/allergies4010002.
2. The ARIA guidelines for classification and treatment of allergic rhinitis, featured in a review article on ScienceDirect. More details can be found by searching the article title on ScienceDirect.
3. Wallace, D. V., Dykewicz, M. S., Oppenheimer, J., Portnoy, J. M., Lang, D. M., & American Academy of Allergy, Asthma & Immunology (AAAAI). (2020). Rhinitis 2020: A practice parameter update. Retrieved from AAAAI website.

These references should provide comprehensive guidelines and research on the management of allergic rhinitis.

9. **Please provide references to scientific papers on guidelines for the benign paroxysmal positional vertigo. Only the bibliographic data of the papers is required.**

Here are several references to scientific papers and guidelines on benign paroxysmal positional vertigo (BPPV) that you might find useful:

1. Bhattacharyya N, Gubbels SP, Schwartz SR, et al. Clinical Practice Guideline: Benign Paroxysmal Positional Vertigo (Update). Otolaryngol Head Neck Surg. 2017;156(3_suppl)

   . DOI: 10.1177/0194599816689667.

2. Fife TD, Iverson DJ, Lempert T, et al. Practice parameter: therapies for benign paroxysmal positional vertigo (an evidence-based review): report of the Quality Standards Subcommittee of the American Academy of Neurology. Neurology. 2008;70(22):2067-2074. DOI: 10.1212/01.wnl.0000313378.77444.ac.

3. Hilton MP, Pinder DK. The Epley (canalith repositioning) manoeuvre for benign paroxysmal positional vertigo. Cochrane Database Syst Rev. 2014;(12)

   . DOI: 10.1002/14651858.CD003162.pub3.

4. von Brevern M, Bertholon P, Brandt T, Fife T, Imai T, Nuti D, Newman-Toker D. Benign paroxysmal positional vertigo: Diagnostic criteria. J Vestib Res. 2015;25(3-4):105-117. DOI: 10.3233/VES-150553.

5. Kim JS, Zee DS. Clinical practice. Benign paroxysmal positional vertigo. N Engl J Med. 2014;370(12):1138-1147. DOI: 10.1056/NEJMcp1309481.

These references include comprehensive reviews and clinical guidelines for the diagnosis, management, and treatment of BPPV. They provide evidence-based recommendations for clinical practice and are recognized by various medical authorities and institutions.

10. **Please provide references to scientific papers on guidelines for the evaluation of hearing preservation in acoustic neuroma. Only the bibliographic data of the papers is required.**

Here is a list of references to scientific papers that focus on guidelines for the evaluation of hearing preservation in acoustic neuroma:

1. Arriaga, M. A., Chen, D. A., & Fukushima, T. (1997). Individualizing hearing preservation in acoustic neuroma surgery. Laryngoscope, 107(8), 1043-1047. doi: 10.1097/00005537-199708000-00007

2. Committee on Hearing and Equilibrium. (1995). Committee on Hearing and Equilibrium guidelines for the evaluation of hearing preservation in acoustic neuroma (vestibular schwannoma). Otolaryngology–Head and Neck Surgery, 113(3), 179-180. doi: 10.1016/S0194-5998(95)70100-4

3. Fukushima, T. (2003). Patient Selection and Method of Evaluation for Hearing Preservation in Acoustic Neuroma Surgery. In Kanzaki, J., Tos, M., Sanna, M., Moffat, D.A., Kunihiro, T., Inoue, Y. (Eds.), Acoustic Neuroma (Vol. 10). Springer, Tokyo. doi: 10.1007/978-4-431-53942-1_17

4. Mazzoni, A., Zanoletti, E., & Denaro, L. (2018). Retrolabyrinthine meatotomy as part of retrosigmoid approach to expose the whole internal auditory canal: rationale, technique and outcome in hearing preservation surgery for vestibular schwannoma. Operative Neurosurgery, 14(1), 36-44. doi: 10.1093/ons/opx034

These papers provide detailed insights into various aspects of hearing preservation strategies in the context of acoustic neuroma treatment, including guidelines for clinical practice, surgical approaches, and outcome evaluations.

**11. Please provide references to scientific papers on guidelines for the otitis media with effusion. Only the bibliographic data of the papers is required.**

Here are some references to scientific papers and guidelines concerning the management of otitis media with effusion (OME):

1. **American Academy of Pediatrics:**

   "Otitis Media With Effusion" by the American Academy of Pediatrics, published in Pediatrics, May 2004, Vol. 113, No. 5, pp. 1412-1429. This article offers guidelines on diagnosing and managing OME in children. View the publication

2. **American Academy of Otolaryngology-Head and Neck Surgery (AAO-HNS):**

   "Clinical Practice Guideline: Otitis Media with Effusion (Update)" by Richard M. Rosenfeld et al., updated in 2016. This guideline provides comprehensive recommendations for the management of OME, emphasizing diagnostic methods, management strategies, and follow-up care. Read more about the guideline

3. **American Academy of Family Physicians (AAFP):**

   This source discusses the updated guidelines released by AAO-HNS for managing OME, highlighting the importance of assessing at-risk children and providing detailed information about the natural history of OME and follow-up needs. Learn more from AAFP

These sources provide a solid foundation of the current guidelines and evidence-based practices for managing OME, particularly in pediatric populations.

**12. Please provide references to scientific papers on guidelines for the Bell's palsy. Only the bibliographic data of the papers is required.**

Here are several references to scientific papers and guidelines concerning Bell's palsy:

1. Baugh RF, Basura GJ, Ishii LE, et al. "Clinical practice guideline: Bell’s palsy." Otolaryngol Head Neck Surg. 2013;149(3_suppl)

   –S27. Available from: doi.org

2. de Almeida JR, Guyatt GH, Sud S, et al. "Management of Bell palsy: clinical practice guideline." CMAJ. 2014;186(12):917–922. Available from: doi.org

3. Gronseth GS, Paduga R. "Evidence-based guideline update: steroids and antivirals for Bell palsy: report of the guideline development subcommittee of the American academy of neurology." Neurology. 2012;79(22):2209–2213. Available from: doi.org

4. Lassaletta L, Morales-Puebla JM, et al. "Facial paralysis: clinical practice guideline of the Spanish society of otolaryngology." Acta Otorrinolaringologica (English Edition). 2020;71(2):99–118. Available from: doi.org

5. Fieux M, Franco-Vidal V, et al. "French society of ENT (SFORL) guidelines. Management of acute Bell's palsy." Eur Ann Otorhinolaryngol Head Neck Dis. 2020;137(6):483–488. Available from: doi.org

These references provide comprehensive information on the diagnosis, management, and treatment of Bell's palsy according to various medical guidelines and expert consensus.

**13. Please provide references to scientific papers on guidelines for the tympanostomy tubes in children. Only the bibliographic data of the papers is required.**

Here are references to some scientific papers and guidelines concerning tympanostomy tubes in children:

1. Rosenfeld RM, Schwartz SR, Pynnonen MA, et al. Clinical Practice Guideline: Tympanostomy Tubes in Children. Otolaryngol Head Neck Surg. 2013 Jul;149(1_suppl)

   . Available from: https://journals.sagepub.com/doi/full/10.1177/0194599813487302

   This guideline addresses patient selection and surgical indications for tympanostomy tubes in children.

2. Rettig E, Tunkel DE. Contemporary Guidelines for Tympanostomy Tube Placement. Curr Treat Options Pediatr. 2020;6:25-36. Available from: https://link.springer.com/article/10.1007/s40746-020-00171-7

   This paper discusses the modern approaches and recommendations for tympanostomy tube placement in pediatric patients.

3. Anne S, Preciado DA, Chelius DC, et al. Clinical Practice Guideline: Tympanostomy Tubes in Children (Update). Otolaryngol Head Neck Surg. 2022 Feb;166(2_suppl)

   . Available from: https://journals.sagepub.com/doi/full/10.1177/01945998221074396

   An updated clinical practice guideline that provides evidence-based recommendations on the management of tympanostomy tubes in children.

These references provide a comprehensive overview of the guidelines and research surrounding the use of tympanostomy tubes in pediatric patients.

**14. Please provide references to scientific papers on guidelines for the tinnitus. Only the bibliographic data of the papers is required.**

Here are some scientific papers providing guidelines for tinnitus management:

**1.** Tinnitus Guidelines and Their Evidence Base

   Authors: Dirk De Ridder, et al.

   Journal: Journal of Clinical Medicine

Publication Date: April 2023

DOI: 10.3390/jcm12093087

2. A Multidisciplinary European Guideline for Tinnitus: Diagnostics, Assessment, and Treatment

   Journal: HNO (No specific authors provided in the snippet)

   Details: This guideline addresses habituation theory and the neurophysiological and cognitive models of tinnitus.

3. Deep Brain Stimulation for Primary Refractory Tinnitus: A Systematic Review

   Authors: Aiyush Bansal, Farrokh Farrokhi

   Journal: Brain Sciences

   Publication Date: April 2024

   DOI: 10.3390/brainsci14050452

4. Clinical Practice Guideline: Tinnitus

   Authors: David E. Tunkel, Carol A. Bauer, et al.

   Journal: Otolaryngology—Head Neck Surgery

   Publication Date: October 2014

   Details: This guideline from the American Academy of Otolaryngology provides recommendations for screening, assessment, and treatment of adults with tinnitus. It distinguishes between bothersome and non-bothersome tinnitus, advises on physical examinations, and suggests interventions like cognitive behavioral therapy and hearing aid evaluations.

These references are a great starting point for understanding the current standards and research in managing tinnitus through evidence-based practices.

**15. Please provide references to scientific papers on guidelines for improving voice outcomes after thyroid surgery. Only the bibliographic data of the papers is required.**

Here are some references to scientific papers that provide guidelines for improving voice outcomes after thyroid surgery:

1. Chandrasekhar SS, Randolph GW, Seidman MD, Rosenfeld RM, et al. Clinical Practice Guideline: Improving Voice Outcomes after Thyroid Surgery. Otolaryngology - Head and Neck Surgery. 2013; 148(6 Suppl): S1-S37.

   This guideline offers comprehensive recommendations for managing voice outcomes in adult patients undergoing thyroid surgery. It emphasizes the importance of identifying the recurrent laryngeal nerve during surgery and includes various recommendations for preoperative and postoperative voice assessments (SpeechBite).

2. Chandrasekhar SS, Randolph GW, Seidman MD, Rosenfeld RM, et al. Clinical Practice Guideline: Improving Voice Outcomes after Thyroid Surgery. Otolaryngology - Head and Neck Surgery. 2013 Jun; 148(6 Suppl): S1-37.

   This document provides evidence-based practices to optimize voice outcomes, focusing on interventions before, during, and after thyroid surgery to manage potential voice issues. The guidelines are targeted at clinicians who interact with thyroid surgery patients, including otolaryngologists, endocrinologists, and speech-language pathologists (ENTnet).

These papers are essential resources for healthcare providers looking to enhance voice care in patients undergoing thyroid procedures, addressing everything from surgical techniques to postoperative care.

**16. Please provide references to scientific papers on guidelines for the hoarseness (dysphonia). Only the bibliographic data of the papers is required.**

Here are some references to scientific papers providing guidelines on hoarseness (dysphonia):

1. Stachler, R. J., Francis, D. O., et al. (2018). Clinical Practice Guideline: Hoarseness (Dysphonia) (Update). Otolaryngology—Head and Neck Surgery, 158(1_Suppl), S1-S42.

This publication from the American Academy of Otolaryngology—Head and Neck Surgery provides updated guidelines on managing hoarseness, aiming to optimize patient care by recommending evidence-based treatments and diagnostic approaches (ASHA).

**17. Please provide references to scientific papers on guidelines for nutritional management in head and neck cancer. Only the bibliographic data of the papers is required.**

Here are several references to scientific papers that focus on guidelines for the nutritional management of head and neck cancer:

1. Arends J., Bachmann P., Baracos V., et al. (2017). ESPEN guidelines on nutrition in cancer patients. Clinical Nutrition, 36, 11-48. DOI: 10.1016/j.clnu.2016.07.015

2. Denaro N., Merlano M.C., Russi E.G. (2013). Dysphagia in head and neck cancer patients: Pretreatment evaluation, predictive factors, and assessment during radio-chemotherapy, recommendations. Clinical and Experimental Otorhinolaryngology, 6(3), 117-126. DOI: 10.3342/ceo.2013.6.3.117

3. Giles K.H., Kubrak C., Baracos V.E., et al. (2016). Recommended European Society of Parenteral and Enteral Nutrition protein and energy intakes and weight loss in patients with head and neck cancer. Head Neck, 38, 1248-1257. DOI: 10.1002/hed.24427

4. Jager-Wittenaar H., Dijkstra P.U., Vissink A., et al. (2011). Changes in nutritional status and dietary intake during and after head and neck cancer treatment. Head Neck, 33, 863-870. DOI: 10.1002/hed.21546

5. Head and Neck Guideline Steering Committee. (2018). Evidence-based practice guidelines for the nutritional management of adult patients with head and neck cancer. Clinical Oncological Society of Australia. Available from COSA
6. Langius J.A.E., Zandbergen M.C., Eerenstein S.E.J., et al. (2013). Effect of nutritional interventions on nutritional status, quality of life and mortality in patients with head and neck cancer receiving (chemo)radiotherapy: a systematic review. Clinical Nutrition, 32(5), 671-678. DOI: 10.1016/j.clnu.2013.06.012
7. Bhattacharjee A., Bahar I., Saikia B.J. (2021). Enteral feeding methods for nutritional management in patients with head and neck cancers being treated with radiotherapy and/or chemotherapy. Cochrane Database of Systematic Reviews. DOI: 10.1002/14651858.cd007904.pub3

These references provide a range of perspectives and recommendations regarding the nutritional management of head and neck cancer patients, including evidence-based guidelines, systematic reviews, and clinical studies focusing on dietary needs, interventions, and outcomes.

**18. Please provide references to scientific papers on guidelines for the acute otitis externa. Only the bibliographic data of the papers is required.**

Here are some references to scientific papers that provide guidelines for the management of acute otitis externa:

1. Rosenfeld, R. M., Schwartz, S. R., Cannon, C. R., Roland, P. S., Simon, G. R., Kumar, K. A., Huang, W. W., Haskell, H. W., & Robertson, P. J. (2014). Clinical Practice Guideline: Acute Otitis Externa (Update). Otolaryngology–Head and Neck Surgery, February 2014 issue. This guideline emphasizes the appropriate use of topical antimicrobials and highlights the need for adequate pain relief, detailing management strategies specifically for diffuse acute otitis externa which is predominantly bacterial.
2. Clinical Practice Guideline: Acute Otitis Externa. AAO-HNS Bulletin. This source provides a comprehensive guideline developed by a multidisciplinary panel, including updated evidence and recommendations for the management of acute otitis externa, commonly known as swimmer's ear.

These references provide a robust framework for clinicians treating acute otitis externa, reflecting the latest clinical trials and systematic reviews to ensure evidence-based practice.

**19. Please provide references to scientific papers on guidelines for management of salivary gland tumours. Only the bibliographic data of the papers is required.**

Here are references to some scientific papers and guidelines on the management of salivary gland tumors:

1. European Society for Medical Oncology (ESMO) Clinical Practice Guidelines for salivary gland cancer, which include recommendations on diagnosis, treatment, and follow-up of salivary gland tumors. These guidelines are comprehensive and cover various aspects including clinical examination, pathology, imaging, and therapeutic approaches for different types of salivary gland tumors.

2. United Kingdom National Multidisciplinary Guidelines on the management of salivary gland tumors, published in "The Journal of Laryngology & Otology." These guidelines provide detailed information on diagnostic techniques like fine needle aspiration and core biopsy, imaging modalities, staging, and specific surgical management strategies for different salivary gland tumors.

3. American Society of Clinical Oncology (ASCO) Guidelines for the management of salivary gland malignancy, which include recommendations on surgical approaches, use of radiation therapy, and considerations for systemic therapy.

These sources offer a wealth of information on current practices and recommendations for managing salivary gland tumors and can serve as valuable references for healthcare professionals involved in the treatment of these conditions. For direct access to these guidelines, you can visit their respective publishers' websites or access them through medical libraries and databases.

**20. Please provide references to scientific papers on guidelines for the peritonsillar abscess. Only the bibliographic data of the papers is required.**

Here are some references to scientific papers that discuss guidelines for peritonsillar abscess:

1. Smith KL, Hughes R, Myrex P. (2023). Tonsillitis and Tonsilloliths: Diagnosis and Management. Am Fam Physician, 107(1), 35-41.

2. Kawabata M, Umakoshi M, Makise T, Miyashita K, Harada M, Nagano H, Ohori J, Kurono Y. (2016). Clinical classification of peritonsillar abscess based on CT and indications for immediate abscess tonsillectomy. Auris Nasus Larynx, 43(2), 182-6. doi: 10.1016/j.anl.2015.09.014.

3. Nguyen T, Haberland CA, Hernandez-Boussard T. (2015). Pediatric Patient and Hospital Characteristics Associated With Treatment of Peritonsillar Abscess and Peritonsillar Cellulitis. Clin Pediatr (Phila), 54(13), 1240-6. doi: 10.1177/0009922814565884.

These references provide insights into the current standards and procedures for managing peritonsillar abscesses.

**21. Please provide references to scientific papers on guidelines for the diagnosis of noise-induced hearing loss. Only the bibliographic data of the papers is required.**

Here are some references on guidelines for the diagnosis of noise-induced hearing loss:

1. Council for Accreditation in Occupational Hearing Conservation (CAOHC) 2019 Update:

   "Occupational noise-induced hearing loss: ACOEM practice guideline," Journal of Occupational and Environmental Medicine, vol. 61, no. 6, June 2019, pp. e267-e280.

   Available at: CAOHC - ACOEM Guideline 2019

2. New Zealand's ACC Guidelines (2018):

"Guidelines for diagnosing occupational noise-induced hearing loss," ACC New Zealand, January 2018.

Available at: ACC New Zealand Guidelines 2018

These documents provide detailed guidelines for assessing and diagnosing noise-induced hearing loss, suitable for medical, legal, and occupational health contexts. They discuss criteria such as hearing threshold levels, audiometric notches, and the impact of occupational noise exposure.

**22. Please provide references to scientific papers on guidelines for the nasal provocations with aspects on nasal patency, airflow, and airflow resistance. Only the bibliographic data of the papers is required.**

Here are some references to scientific papers that provide guidelines for nasal provocations with aspects on nasal patency, airflow, and airflow resistance:

1. Malm, L., Van Wijk, R. G., & Bachert, C. (2000). Guidelines for nasal provocations with aspects on nasal patency, airflow, and airflow resistance. Rhinology, 38(1), 1–6.

   This paper discusses consensus efforts on the indications and techniques for nasal provocation and critically analyzes methods to measure their effects, emphasizing the clinical and research applications of techniques such as rhinomanometry and acoustic rhinometry.

2. Walden, S. M., Proud, D., Bascom, R., et al. (1988). Experimentally induced nasal allergic responses. Journal of Allergy and Clinical Immunology, 81, 940–949.

   This research elaborates on methods used for experimentally inducing nasal allergic responses, which could provide insights into the protocols and evaluations used in nasal provocations.

3. Sicherer, S., Wood, R. A., & Eggleston, P. A. (1997). Determinants of airway responses to cat allergen: Comparison of environmental challenge to quantitative nasal and bronchial allergen challenge. Journal of Allergy and Clinical Immunology, 99, 798–805.

   This study compares environmental allergen challenges with quantitative nasal and bronchial provocations, offering a detailed view on airway responses that might be beneficial for understanding nasal patency dynamics.

These articles should provide comprehensive insights and guidelines into the methods and evaluations associated with nasal provocations concerning nasal patency, airflow, and resistance.

**23. Please provide references to scientific papers on guidelines for the school-based bilingual assessment. Only the bibliographic data of the papers is required.**

Here are references to scientific papers on school-based bilingual assessment, focusing on guidelines and practices:

1. Caesar, L. G., & Kohler, P. D. (2007). The State of School-Based Bilingual Assessment: Actual Practice Versus Recommended Guidelines. Language, Speech, and Hearing Services in Schools, 38(3), 190-200. DOI: 10.1044/0161-1461(2007/020)
2. Bonifacci, P., Atti, E., Casamenti, M., Piani, B., Porrelli, M., & Mari, R. (2020). Which measures better discriminate language minority bilingual children with and without developmental language disorder? A study testing a combined protocol of first and second language assessment. Journal of Speech, Language, and Hearing Research, 63(6), 1898–1915. DOI: 10.1044/2020_JSLHR-19-00385
3. Castilla-Earls, A., Pérez-Leroux, A. T., Fulcher-Rood, K., Barr, C. (2021). Morphological errors in Spanish-speaking bilingual children with and without developmental language disorders. Language, Speech, and Hearing Services in Schools, 52(2), 497–511. DOI: 10.1044/2020_LSHSS-19-00073

These papers discuss the practical application of assessment guidelines, the effectiveness of bilingual assessment protocols, and common errors in assessments, providing a comprehensive overview of current practices and recommendations in the field of school-based bilingual assessment.

**24. Please provide references to scientific papers on guidelines for the acquired apraxia of speech. Only the bibliographic data of the papers is required.**

Here are some references to scientific papers on guidelines for acquired apraxia of speech:

1. Wambaugh, J. L., Duffy, J. R., McNeil, M. R., Robin, D. A., & Rogers, M. A. (2006). Treatment Guidelines for Acquired Apraxia of Speech: A Synthesis and Evaluation of the Evidence. Journal of Medical Speech-Language Pathology, 14(2), 15-33. This paper is a systematic review that critically appraises the evidence utilized in developing treatment guidelines for acquired apraxia of speech, covering fifty-nine publications and evaluating them across 33 variables.
2. Ballard, K. J., Wambaugh, J. L., et al. (2015). Treatment for Acquired Apraxia of Speech: A Systematic Review of Intervention Research Between 2004 and 2012. American Journal of Speech-Language Pathology, 24(2), 316-337. This review focuses on the effects of various treatment approaches on impairment or function in individuals with acquired apraxia of speech, examining studies published over an eight-year period.

These articles offer comprehensive reviews of the existing research on treatment approaches and guidelines for acquired apraxia of speech, providing valuable insights for both clinical application and further research.

**25. Please provide references to scientific papers on guidelines for the safe tracheostomy during the COVID-19 pandemic. Only the bibliographic data of the papers is required.**

Here are some references to scientific papers providing guidelines for safe tracheostomy procedures during the COVID-19 pandemic:

1. McGrath, B.A., Brenner, M.J., Warrillow, S.J., Pandian, V., Arora, A., Cameron, T.S., ... & Higgs, A. (2020). Tracheostomy in the COVID-19 era: global and

multidisciplinary guidance. The Lancet Respiratory Medicine, 8(7), 717-725. DOI: 10.1016/S2213-2600(20)30230-7

2. Takhar, A., Walker, A., Tricklebank, S., Wyncoll, D., Hart, N., Jacob, T., ... & McGrath, B. A. (2020). Recommendation of a practical guideline for safe tracheostomy during the COVID-19 pandemic. European Archives of Oto-Rhino-Laryngology, 277, 2173–2184. DOI: 10.1007/s00405-020-06002-z
3. Queen, E. A., & van der Merwe, M. (2020). Early Outcomes From Early Tracheostomy for Patients With COVID-19: A Systematic Review and Meta-analysis. JAMA Otolaryngology–Head & Neck Surgery, 146(9), 850-854. DOI: 10.1001/jamaoto.2020.1770
4. Andriolo, B. N., Andriolo, R. B., Saconato, H., Atallah, Á. N., & Valente, O. (2021). Early versus late tracheostomy for critically ill patients. Cochrane Database of Systematic Reviews, 1(1), CD007271. DOI: 10.1002/14651858.CD007271.pub4
5. Okusanya, K. A., Palmieri, P. A., McGrath, B. A., Pelosi, P., & Sole, M. L. (2020). Critical Care Guidance for Tracheostomy Care During the COVID-19 Pandemic: A Global, Multidisciplinary Approach. American Journal of Critical Care, 29(6), e116-e127. DOI: 10.4037/ajcc2028784

These references include a range of guidelines and recommendations, discussing the timing of tracheostomy, procedural modifications to reduce risk, and management after the procedure. Each of these papers should provide a comprehensive understanding of the safety measures and protocols recommended during the pandemic.

### ChatGPT-4 responses - 10.07.2024

1. **Please provide references to scientific papers on guidelines for the diagnosis and evaluation of therapy in Meniere’s-disease. Only the bibliographic data of the papers is required.**

Here are some references to scientific papers that discuss guidelines for the diagnosis and evaluation of therapy in Meniere's disease:

1. Basura, G. J., Adams, M. E., et al. (2020). Clinical Practice Guideline: Ménière’s Disease. Otolaryngology–Head and Neck Surgery, 162(2 Suppl), S1-S55. This guideline provides comprehensive recommendations for healthcare providers on diagnosing and managing Meniere's disease in adults.

2. Committee on Hearing and Equilibrium guidelines for the diagnosis and evaluation of therapy in Meniere’s disease. American Academy of Otolaryngology-Head and Neck Foundation, Inc. Otolaryngol Head Neck Surg. 1995;113(3):181–5. This older guideline offers foundational insights into the diagnosis and therapeutic evaluations for Meniere's disease.

These resources can serve as a solid starting point for understanding the current clinical approaches to diagnosing and treating Meniere’s disease.

2. **Please provide references to scientific papers on guidelines for the adult sinusitis. Only the bibliographic data of the papers is required.**

Here are some references to scientific papers that include guidelines for managing adult sinusitis:

1. Rosenfeld, R. M., Piccirillo, J. F., Chandrasekhar, S. S., Brook, I., Kumar, K. A., Kramper, M., ... & Corrigan, M. D. (2015). Clinical Practice Guideline (Update): Adult Sinusitis. Otolaryngology–Head and Neck Surgery, 152(2S), S1-S39. This guideline identifies quality improvement opportunities in managing adult rhinosinusitis and provides explicit recommendations for clinical practice.

2. Rosenfeld, R. M., Piccirillo, J. F., Chandrasekhar, S. S., Brook, I., Ashok Kumar, K., Kramper, M., ... & Walsh, S. A. (2015). Updated Clinical Practice Guideline: Adult Sinusitis. American Academy of Otolaryngology-Head and Neck Surgery Bulletin. This update emphasizes shared decision-making and judicious use of antibiotics, highlighting the use of amoxicillin with or without clavulanate when antibiotics are prescribed.

These guidelines cover a range of topics including the differentiation between viral and bacterial sinusitis, appropriate use of antibiotics, and the importance of imaging and other diagnostic tests in the management of sinusitis. They are intended for clinicians managing adults with rhinosinusitis across various settings.

3. **Please provide references to scientific papers on guidelines for the sudden hearing loss. Only the bibliographic data of the papers is required.**

Here are some references to scientific papers on guidelines for sudden hearing loss:

1. Chandrasekhar, S. S., Tsai Do, B., et al. (2019). Clinical Practice Guideline: Sudden Hearing Loss (Update). Otolaryngology–Head and Neck Surgery, 161(1 Suppl), S1-S45. This guideline is updated from its 2012 version and provides comprehensive evidence-based recommendations for managing sudden sensorineural hearing loss.

You can find more detailed recommendations and procedural guidelines in these publications to assist healthcare professionals in the prompt recognition and management of sudden hearing loss.

4. **Please provide references to scientific papers on guidelines for the functional assessment of voice pathology. Only the bibliographic data of the papers is required.**

Here are some references to scientific papers that provide guidelines for the functional assessment of voice pathology:

These papers cover various aspects of voice pathology assessment, including telehealth applications, treatment outcomes, and validity studies of specific assessment scales.

5. **Please provide references to scientific papers on guidelines for the electrophysiologic recurrent laryngeal nerve monitoring during thyroid surgery. Only the bibliographic data of the papers is required.**

Here are some scientific papers with guidelines on electrophysiological recurrent laryngeal nerve monitoring during thyroid surgery:

1. Randolph, G. W., Dralle, H., Abdullah, H., et al. (2011). Electrophysiologic Recurrent Laryngeal Nerve Monitoring During Thyroid and Parathyroid Surgery: International Standards Guideline Statement. Laryngoscope, 121 Suppl 1, S1–S16. DOI: 10.1002/lary.21119.

2. Barczynski, M., Randolph, G. W., Cernea, C. R., Dralle, H., Dionigi, G., Alesina, P. F., et al. (2013). External Branch of the Superior Laryngeal Nerve Monitoring During Thyroid and Parathyroid Surgery: International Neural Monitoring Study Group Standards Guideline Statement. Laryngoscope, 123 Suppl 4, S1–S14. DOI: 10.1002/lary.24301.

3. Wu, C. W., Dionigi, G., Barczynski, M., Chiang, F. Y., Dralle, H., Schneider, R., et al. (2018). International Neuromonitoring Study Group Guidelines 2018: Part II: Optimal Recurrent Laryngeal Nerve Management for Invasive Thyroid Cancer-Incorporation of Surgical, Laryngeal, and Neural Electrophysiologic Data. Laryngoscope, 128 Suppl 3, S18–S27. DOI: 10.1002/lary.27360.

These guidelines provide detailed protocols for intraoperative nerve monitoring to improve surgical outcomes and reduce complications related to nerve damage during thyroid surgeries. They cover various aspects of monitoring techniques, including both intermittent and continuous monitoring approaches, and integrate data from different domains to enhance decision-making during surgery.

**6. Please provide references to scientific papers on guidelines for the tonsillectomy in children. Only the bibliographic data of the papers is required.**

Here are some references to scientific papers and guidelines on tonsillectomy in children:

1. Mitchell, R. B., Archer, S. M., Ishman, S. L., Rosenfeld, R. M., Coles, S., Finestone, S. A., ... & Nnacheta, L. (2019). Clinical Practice Guideline: Tonsillectomy in Children (Update). Otolaryngology–Head and Neck Surgery, 160(2_suppl), S1-S42. DOI: 10.1177/0194599818801757

2. American Academy of Otolaryngology–Head and Neck Surgery Foundation. (2019). Clinical Practice Guideline: Tonsillectomy in Children (Update). Available at: https://effectivehealthcare.ahrq.gov/sites/default/files/pdf/aao-guidelines-tonsillectomy-2019.pdf

These papers provide updated, evidence-based recommendations on the pre-, intra-, and postoperative care and management of children aged 1 to 18 years who are under consideration for tonsillectomy. They are intended for all clinicians in any setting who interact with children who may be candidates for tonsillectomy.

**7. Please provide references to scientific papers on guidelines for the evaluation of results of treatment of conductive hearing-loss. Only the bibliographic data of the papers is required.**

Here are some scientific papers on guidelines for evaluating the results of treatment for conductive hearing loss, with their bibliographic information:

1. Barbara, M., Perotti, M., Gioia, B., Volpini, L., Monini, S. (2020). "Effectiveness of Bonebridge system in patients with conductive hearing loss: A multi-center study". European Archives of Oto-Rhino-Laryngology. This study evaluates the Bonebridge BCI 602 implant for adults with conductive or mixed hearing loss, focusing on audiological outcomes and patient satisfaction (SpringerLink).

2. Pichora-Fuller, M.K., Dupuis, K., Reed, M., Lemke, U. (2013). "Hearing loss and cognitive decline in older adults: Are they connected?". Journal of Speech, Language, and Hearing Research. This paper discusses the relationship between hearing loss, including conductive types, and cognitive decline, providing insights into the broader implications of auditory health (ASHA).

3. National Institute for Health and Care Excellence (NICE). (2023). "Hearing loss in adults: Assessment and management". NICE guideline [NG98]. This guideline provides comprehensive protocols for assessing and managing hearing loss in adults within various healthcare settings, updated as of October 2023 (NICE).

These references should provide a solid foundation for understanding current methodologies and outcomes related to the treatment of conductive hearing loss.

8. **Please provide references to scientific papers on guidelines for the allergic rhinitis. Only the bibliographic data of the papers is required.**

Here are some references to scientific papers on guidelines for allergic rhinitis:

1. Klimek, L., Pfaar, O., Worm, M., Eiwegger, T., Hagemann, J., Ollert, M., ... & Wagenmann, M. (2019). ARIA guideline 2019: Treatment of allergic rhinitis in the German health system. Allergo Journal International. This publication provides a summary of the guidelines developed after the ARIA (Allergic Rhinitis and its Impact on Asthma) initiative's meeting in Paris, adapted for the German health system.

2. Dykewicz, M. S., Wallace, D. V., Baroody, F., Bernstein, J., Craig, T., Finegold, I., ... & Blessing-Moore, J. (2020). Rhinitis 2020: A practice parameter update. This document provides comprehensive practice parameters for allergic rhinitis and nonallergic rhinitis, covering diagnosis, assessment, and treatment strategies.

These papers offer extensive guidance on managing allergic rhinitis based on recent research and consensus among experts in the field. For further details, you might access these documents through academic libraries or contact the respective publishers.

9. **Please provide references to scientific papers on guidelines for the benign paroxysmal positional vertigo. Only the bibliographic data of the papers is required.**

Here are the bibliographic references for guidelines on Benign Paroxysmal Positional Vertigo (BPPV) from reputable sources:

1. American Academy of Otolaryngology—Head and Neck Surgery (AAO-HNS): Bhattacharyya, N., Gubbels, S.P., Schwartz, S.R., et al. Clinical Practice Guideline: Benign Paroxysmal Positional Vertigo (Update). Otolaryngology–Head and Neck Surgery, March 1, 2017. Available at: AAO-HNS Journals.

2. American Academy of Audiology: Clinical Practice Guideline: Benign Paroxysmal Positional Vertigo (Update). Published on March 1, 2017, and endorsed by various organizations including the American Academy of Physical Medicine and Rehabilitation and the American Neurotology Society. Accessible at: American Academy of Audiology.

These guidelines are aimed at improving the management of BPPV by promoting accurate diagnosis, reducing the use of unnecessary treatments, and enhancing patient care and outcomes. They provide valuable insights for clinicians involved in the diagnosis and management of BPPV.

**10. Please provide references to scientific papers on guidelines for the evaluation of hearing preservation in acoustic neuroma. Only the bibliographic data of the papers is required.**

Here are some key references regarding guidelines for the evaluation of hearing preservation in acoustic neuroma:

1. Committee on Hearing and Equilibrium (1995)**:** This foundational paper outlines the guidelines for the evaluation of hearing preservation in patients with acoustic neuroma, also known as vestibular schwannoma. Published in Otolaryngology–Head and Neck Surgery, this is a seminal work in the field.

   Citation: Committee on Hearing and Equilibrium. "Guidelines for the evaluation of hearing preservation in acoustic neuroma (vestibular schwannoma)." Otolaryngol Head Neck Surg 113, no. 179-180 (1995).

2. Arriaga, M.A., Chen, D.A., and Fukushima, T. (1997)**:** This paper discusses individualized approaches to preserving hearing in acoustic neuroma surgery, emphasizing tailored treatment plans based on patient-specific factors.

   Citation: Arriaga, M.A., Chen, D.A., Fukushima, T. "Individualizing hearing preservation in acoustic neuroma surgery." Laryngoscope 107, no. 1043-1047 (1997).

3. Fukushima, T. (2003): In a book chapter from the proceedings of the Keio University International Symposia for Life Sciences and Medicine, Fukushima explores patient selection and evaluation methods for hearing preservation in acoustic neuroma surgery, providing a comprehensive look at the decision-making process and surgical techniques.

   Citation: Fukushima, T. "Patient Selection and Method of Evaluation for Hearing Preservation in Acoustic Neuroma Surgery." In: Kanzaki, J., Tos, M., Sanna, M., Moffat, D.A., Kunihiro, T., Inoue, Y. (eds) Acoustic Neuroma. Keio University International Symposia for Life Sciences and Medicine, vol 10. Springer, Tokyo. https://doi.org/10.1007/978-4-431-53942-1_17 (2003).

These papers collectively provide a strong basis for understanding current practices and recommendations in the management of hearing preservation during the treatment of acoustic neuroma. For detailed information and access to these studies, you might need to check with institutional libraries or purchase access through academic journals.

**11. Please provide references to scientific papers on guidelines for the otitis media with effusion. Only the bibliographic data of the papers is required.**

Here are some references to scientific papers and guidelines concerning otitis media with effusion:

1. Rosenfeld, R. M., Shin, J. J., Schwartz, S. R., Coggins, R., Gagnon, L., Hackell, J. M., ... & Vila, P. M. (2016). Clinical Practice Guideline: Otitis Media with Effusion (Update). Otolaryngology–Head and Neck Surgery, 154(2), 201-214. This guideline

offers comprehensive recommendations for the diagnosis and management of otitis media with effusion (OME) in children (EntNet).

2. American Academy of Pediatrics, Subcommittee on Otitis Media With Effusion, & American Academy of Family Physicians. (2004). Otitis Media With Effusion. Pediatrics, 113(5), 1412-1429. This article from Pediatrics journal provides evidence-based recommendations on diagnosing and managing OME in children (AAP Publications).

3. Rosenfeld, R. M., Culpepper, L., Doyle, K. J., Grundfast, K. M., Hoberman, A., Kenna, M. A., ... & Yawn, B. P. (2004). Clinical Practice Guideline: Otitis Media with Effusion. Otolaryngology–Head and Neck Surgery, 130(Suppl 5), S95-S118. This guideline covers the management of OME and highlights the need for accurate diagnosis and effective communication between clinicians and patients (EntNet).

These references provide valuable guidelines and insights into the management of otitis media with effusion, based on the latest research and expert consensus.

**12. Please provide references to scientific papers on guidelines for the Bell's palsy. Only the bibliographic data of the papers is required.**

Here are some references to scientific papers that provide guidelines for the management of Bell's palsy:

1. Baugh RF, Basura GJ, Ishii LE, Schwartz SR, Drumheller CM, Burkholder R, Deckard NA, Dawson C, Driscoll C, Gillespie MB, et al. (2013). "Clinical practice guideline: Bell's palsy." Otolaryngol Head Neck Surg, 149(3_suppl)

   –S27.

2. Gronseth GS, Paduga R. (2012). "Evidence-based guideline update: steroids and antivirals for Bell palsy: report of the guideline development subcommittee of the American academy of neurology." Neurology, 79(22):2209–2213.

3. de Almeida JR, Guyatt GH, Sud S, Dorion J, Hill MD, Kolber MR, Lea J, Reg SL, Somogyi BK, Westerberg BD. (2014). "Management of Bell palsy: clinical practice guideline." CMAJ, 186(12):917–922.

4. Lassaletta L, Morales-Puebla JM, Altuna X, Arbizu Á, Arístegui M, Batuecas Á, Cenjor C, Espinosa-Sánchez JM, García-Iza L, García-Raya P, et al. (2020). "Facial paralysis: clinical practice guideline of the Spanish society of otolaryngology." Acta Otorrinolaringologica (English Edition), 71(2):99–118.

5. Fieux M, Franco-Vidal V, Devic P, Bricaire F, Charpiot A, Darrouzet V, Denoix L, Gatignol P, Guevara N, Montava M, Roch JA, et al. (2020). "French society of ENT (SFORL) guidelines. Management of acute Bell's palsy." Eur Ann Otorhinolaryngol Head Neck Dis, 137(6):483–488.

These references encompass a range of international guidelines and are sourced from established journals in the field of otolaryngology and neurology, reflecting a broad consensus on the management of Bell's palsy.

**13. Please provide references to scientific papers on guidelines for the tympanostomy tubes in children. Only the bibliographic data of the papers is required.**

Here are some references to scientific papers and guidelines concerning tympanostomy tubes in children, focusing on the bibliographic details:

1. Rosenfeld, R. M., Schwartz, S. R., Pynnonen, M. A., Tunkel, D. E., Hussey, H. M., Fichera, J. S., ... & Vaughn, W. (2013). Clinical Practice Guideline: Tympanostomy Tubes in Children. Otolaryngology–Head and Neck Surgery, 149(1_suppl), S1-S35. Published July 1, 2013. DOI: 10.1177/0194599813487302. This guideline provides evidence-based recommendations on patient selection and surgical indications for managing tympanostomy tubes in children.

2. Rosenfeld, R. M., Shin, J. J., Schwartz, S. R., Coggins, R., Gagnon, L., Hackell, J. M., ... & Walsh, S. (2016). Clinical Practice Guideline: Otitis Media with Effusion (Update). Otolaryngology–Head and Neck Surgery, 154(1_suppl), S1-S41. Published February 1, 2016. DOI: 10.1177/0194599815623467. This updated guideline addresses comprehensive management strategies for otitis media with effusion, which often precedes the decision for tympanostomy tubes.

3. American Academy of Otolaryngology-Head and Neck Surgery. (2022). Clinical Practice Guideline: Tympanostomy Tubes in Children (Update). Otolaryngology–Head and Neck Surgery. Published February 9, 2022. This update reassesses and provides updated evidence-based recommendations, including patient selection and management nuances for tympanostomy tubes in children. Endorsed by the American Academy of Audiology and other relevant professional groups.

These references should provide a solid basis for exploring the current guidelines and evidence-based practices in the management of tympanostomy tubes in children.

**14. Please provide references to scientific papers on guidelines for the tinnitus. Only the bibliographic data of the papers is required.**

Here are some bibliographic references to scientific papers and guidelines on tinnitus:

1. J. Clin. Med. "Tinnitus Guidelines and Their Evidence Base." 2023. This paper discusses evidence-based medicine as it applies to tinnitus guidelines, covering the formulation of clinical recommendations based on systematic reviews and meta-analyses. Link to article

2. HNO. "A multidisciplinary European guideline for tinnitus: diagnostics, assessment, and treatment." This guideline aims to standardize the diagnosis and treatment of tinnitus across Europe. Link to article

3. Annals of Behavioral Medicine. "The efficacy of auditory perceptual training for tinnitus: a systematic review." 2010. This systematic review by Hoare DJ et al. explores the effectiveness of auditory training in tinnitus management. Link to article

4. American Journal of Otolaryngology. "Efficacy of tinnitus retraining therapy: a meta-analysis and systematic review." 2021. This meta-analysis by Han M, Yang X, Lv J, reviews the outcomes of tinnitus retraining therapy. Link to article

5. BMJ. "Assessment and management of tinnitus: summary of NICE guidance." 2020. This guideline provides a comprehensive overview of the recommended approaches for managing tinnitus as suggested by NICE. Link to article

6. Otolaryngology–Head and Neck Surgery. "Clinical Practice Guideline: Tinnitus." 2014. Developed by the American Academy of Otolaryngology—Head and Neck Surgery, this guideline offers detailed evidence-based recommendations for tinnitus management. Link to article

These references provide a solid starting point for a comprehensive understanding of current guidelines and research concerning the management of tinnitus.

**15. Please provide references to scientific papers on guidelines for improving voice outcomes after thyroid surgery. Only the bibliographic data of the papers is required.**

Here are several scientific papers that discuss guidelines and techniques for improving voice outcomes after thyroid surgery:

1. Wang, J. J., Huang, T. Y., Wu, C. W., et al. (2022). Improving Voice Outcomes After Thyroid Surgery – Review of Safety Parameters for Using Energy-Based Devices Near the Recurrent Laryngeal Nerve. Frontiers in Surgery. This paper discusses the use of energy-based devices in thyroid surgery and their effects on the recurrent laryngeal nerve, which is crucial for voice outcomes. Available at Frontiers in Surgery.

2. Pace-Asciak, P., Russell, J. O., Tufano, R. P. (2022). Improving Voice Outcomes after Thyroid Surgery and Ultrasound-Guided Ablation Procedures. Frontiers in Surgery. This article reviews both surgical and non-surgical approaches to minimize voice complications post-thyroidectomy. Available at Frontiers in Surgery.

3. Chandrasekhar, S. S., Randolph, G. W., Seidman, M. D., et al. (2013). Clinical Practice Guideline: Improving Voice Outcomes after Thyroid Surgery. Otolaryngology–Head and Neck Surgery. This guideline offers evidence-based recommendations to optimize voice outcomes, covering preoperative, intraoperative, and postoperative periods. Available at Otolaryngology–Head and Neck Surgery.

These papers provide a comprehensive overview of current strategies and guidelines for minimizing voice disturbances following thyroid operations, focusing on both technological advancements and clinical practices.

16. **Please provide references to scientific papers on guidelines for the hoarseness (dysphonia). Only the bibliographic data of the papers is required.**

Here are some bibliographic references to scientific papers on guidelines for hoarseness (dysphonia):

1. Stachler, R. J., Francis, D. O., et al. (2018). "Clinical Practice Guideline: Hoarseness (Dysphonia) (Update)." Otolaryngology—Head and Neck Surgery, 158(1_Suppl), S1-S42. This guideline is updated by the American Academy of Otolaryngology—Head and Neck Surgery and provides comprehensive recommendations for managing dysphonia (ASHA).

2. Stachler, R. J., Francis, D. O., Schwartz, S. R., et al. (2018). Published in Otolaryngology—Head and Neck Surgery on March 1, 2018. This publication offers updated evidence-based guidelines for the treatment and assessment of hoarseness, aiming to standardize care and improve patient outcomes (EntNet).

These references are from established medical journals and provide a solid foundation for understanding the current guidelines in the treatment and management of hoarseness or dysphonia.

17. **Please provide references to scientific papers on guidelines for nutritional management in head and neck cancer. Only the bibliographic data of the papers is required.**

Here are several references for scientific papers on guidelines for the nutritional management of head and neck cancer:

1. Arends, J., Bodoky, G., Bozzetti, F., Fearon, K., Muscaritoli, M., Selga, G., ... & Zander, A. (2017). ESPEN guidelines on nutrition in cancer patients. Clinical Nutrition, 36(1), 11-48. This guideline provides comprehensive recommendations for the nutritional management of cancer patients, including those with head and neck cancer. Link to publication

2. Silander, E., Nyman, J., Hammerlid, E. (2013). An overview of the European Society for Clinical Nutrition and Metabolism (ESPEN) guidelines for the nutritional management of patients with head and neck cancer. Dysphagia, 28(2), 264-274. This paper discusses ESPEN's guidelines focusing on strategies to manage the nutritional needs of head and neck cancer patients. Link to publication

3. Findlay, M., Bauer, J., & Brown, T. (2020). Australian guidelines for the nutritional management of adult patients with head and neck cancer. Journal of Human Nutrition and Dietetics, 33(3), 357-367. These guidelines outline best practices for assessing and managing nutritional issues in head and neck cancer patients in the Australian context. Link to publication

4. Langius, J. A., Zandbergen, M. C., Eerenstein, S. E., van Tulder, M. W., Leemans, C. R., Kramer, M. H., & Weijs, P. J. (2013). Effect of nutritional interventions on nutritional status, quality of life and mortality in patients with head and neck cancer receiving (chemo)radiotherapy: A systematic review. Clinical Nutrition, 32(5), 671-

678. This review evaluates the impact of nutritional interventions on outcomes in head and neck cancer patients undergoing treatment. Link to publication

These references offer a range of perspectives and data regarding the nutritional management of head and neck cancer, from general guidelines to specific interventions and their effects.

**18. Please provide references to scientific papers on guidelines for the acute otitis externa. Only the bibliographic data of the papers is required.**

Here are some references to scientific papers on guidelines for acute otitis externa:

1. Rosenfeld, R. M., Schwartz, S. R., Cannon, C. R., et al. (2014). Clinical Practice Guideline: Acute Otitis Externa. Otolaryngology—Head and Neck Surgery, 150(1_suppl), S1-S24. DOI: 10.1177/0194599813517083.

2. Roland, P. S., Stroman, D. W. (2002). Microbiology of Acute Otitis Externa. The Laryngoscope, 112(7), 1166-1177. DOI: 10.1097/00005537-200207000-00010.

3. Dibb, W. L. (1991). Microbial Aetiology of Otitis Externa. Journal of Infection, 22(3), 233-239. DOI: 10.1016/S0163-4453(91)91386-4.

These references include comprehensive guidelines and studies on the microbial causes and management strategies for acute otitis externa.

**19. Please provide references to scientific papers on guidelines for management of salivary gland tumours. Only the bibliographic data of the papers is required.**

Here are some references to scientific papers on the guidelines for the management of salivary gland tumors:

1. van Herpen C, Vander Poorten V, Skalova A, et al. Salivary gland cancer: ESMO–EURACAN Clinical Practice Guideline for diagnosis, treatment and follow-up. Ann Oncol. 2021;32(4):469-489. doi:10.1016/j.annonc.2021.02.019.

2. Sood S, McGurk M, Vaz F. Management of Salivary Gland Tumours: United Kingdom National Multidisciplinary Guidelines. J Laryngol Otol. 2016;130(S2)

   . doi:10.1017/S0022215116000527.

These sources provide comprehensive guidelines for the management of salivary gland tumors, including diagnostic, treatment, and follow-up recommendations.

**20. Please provide references to scientific papers on guidelines for the peritonsillar abscess. Only the bibliographic data of the papers is required.**

Here are some references to scientific papers providing guidelines and information on the management of peritonsillar abscess:

1. Klug TE, Greve T, Hentze M. "Complications of peritonsillar abscess." Ann Clin Microbiol Antimicrob. 2020 Jul 30;19(1):32. doi: 10.1186/s12941-020-00375-x. PMID: 32731900. This paper discusses the complications associated with peritonsillar abscesses.

2. Hur K, Zhou S, Kysh L. "Adjunct steroids in the treatment of peritonsillar abscess: A systematic review." Laryngoscope. 2018 Jan;128(1):72-77. doi: 10.1002/lary.26672. PMID: 28561258. This systematic review evaluates the use of steroids as an adjunct treatment for peritonsillar abscess.

3. Sievert M, Miksch M, Mantsopoulos K, et al. "The value of transcutaneous ultrasound in the diagnosis of tonsillar abscess: A retrospective analysis." Auris Nasus Larynx. 2021 Dec;48(6):1120-1125. doi: 10.1016/j.anl.2021.04.006. PMID: 33906745. This study assesses the effectiveness of ultrasound in diagnosing tonsillar abscesses.

These papers can provide detailed insights into the management practices, complications, and diagnostic techniques for peritonsillar abscesses.

**21. Please provide references to scientific papers on guidelines for the diagnosis of noise-induced hearing loss. Only the bibliographic data of the papers is required.**

Here are some references to scientific papers that provide guidelines for diagnosing noise-induced hearing loss:

1. Coles, R.R.A., Lutman, M.E., Buffin, J.T. (2000). Guidelines on the diagnosis of noise-induced hearing loss for medicolegal purposes. Clinical Otolaryngology and Allied Sciences, 25(4), 264-273. DOI: 10.1046/j.1365-2273.2000.00368.x.

2. Coles, R.R.A., Lutman, M.E., Buffin, J.T. (2000). Guidelines on the diagnosis of noise-induced hearing loss for medicolegal purposes. Clinical Otolaryngology, DOI: 10.1046/j.1365-2273.2000.00368.x.

These articles discuss the criteria and considerations necessary for the medical-legal diagnosis of noise-induced hearing loss, including the identification of high-frequency hearing impairment, exposure to noise, and audiometric notches or bulges. They also consider other modifying factors like age and other causes of hearing loss.

**22. Please provide references to scientific papers on guidelines for the nasal provocations with aspects on nasal patency, airflow, and airflow resistance. Only the bibliographic data of the papers is required.**

Here are references to scientific papers that provide guidelines and examine aspects of nasal patency, airflow, and airflow resistance in the context of nasal provocations:

1. Malm, L., Van Wijk, R. G., & Bachert, C. (2000). Guidelines for nasal provocations with aspects on nasal patency, airflow, and airflow resistance. Rhinology, 38(1), 1–6. This paper discusses the consensus on indications and techniques for nasal

provocation and reviews methods like rhinomanometry and acoustic rhinometry for measuring nasal patency and airflow resistance.

2. Airaksinen, L., Tuomi, T., Vanhanen, M., Voutilainen, R., & Toskala, E. (2007). Use of nasal provocation test in the diagnostics of occupational rhinitis. Rhinology, 45, 40–46. This study focuses on the use of nasal provocation tests to diagnose occupational rhinitis, exploring methods for assessing nasal airflow and resistance.

3. Sicherer, S., Wood, R. A., & Eggleston, P. A. (1997). Determinants of airway responses to cat allergen: Comparison of environmental challenge to quantitative nasal and bronchial allergen challenge. Journal of Allergy and Clinical Immunology, 99, 798–805. This research compares nasal and bronchial allergen challenges to evaluate airway responses, which is vital in understanding the mechanisms affecting nasal patency and airflow during provocation tests.

These references should be useful for a deeper understanding of methodologies and guidelines concerning nasal provocations related to nasal airflow and resistance.

**23. Please provide references to scientific papers on guidelines for the school-based bilingual assessment. Only the bibliographic data of the papers is required.**

Here are some references to scientific papers on guidelines for school-based bilingual assessment:

1. Caesar, Lena G., and Paula D. Kohler. "The State of School-Based Bilingual Assessment: Actual Practice Versus Recommended Guidelines." Language, Speech, and Hearing Services in Schools, vol. 38, no. 3, 2007, pp. 190-200. DOI: 10.1044/0161-1461(2007/020).

2. Gillam, R. B., Peña, E. D., & Miller, L. "Dynamic assessment of narrative and expository discourse." Topics in Language Disorders, vol. 20, 1999, pp. 33–47. DOI: 10.1097/00011363-199911000-00005.

3. Castilla-Earls, A., Pérez-Leroux, A. T., Fulcher-Rood, K., Barr, C. "Morphological errors in Spanish-speaking bilingual children with and without developmental language disorders." Language, Speech, and Hearing Services in Schools, vol. 52, no. 2, 2021, pp. 497–511.

These papers explore various aspects of bilingual assessment in school settings, including the practical application of guidelines and the assessment of specific language impairments in bilingual children.

**24. Please provide references to scientific papers on guidelines for the acquired apraxia of speech. Only the bibliographic data of the papers is required.**

Here are some references for scientific papers on guidelines for acquired apraxia of speech:

1. Wambaugh, J. L., Duffy, J. R., McNeil, M. R., Robin, D. A., & Rogers, M. A. (2006). Treatment Guidelines for Acquired Apraxia of Speech: A Synthesis and Evaluation of the Evidence. Journal of Medical Speech Language Pathology, 14(2), 15-33. This paper presents a systematic review and critical appraisal of treatment guidelines for acquired apraxia of speech.

2. Ballard, K. J., Wambaugh, J. L., et al. (2015). Treatment for Acquired Apraxia of Speech: A Systematic Review of Intervention Research Between 2004 and 2012. American Journal of Speech-Language Pathology, 24(2), 316-337. This review investigates various treatment approaches for apraxia of speech.

3. Munasinghe, T. U., Ariyasena, A. D. K., et al. (2023). Speech Therapy Interventions for Acquired Apraxia of Speech: An Updated Systematic Review. American Journal of Speech-Language Pathology, 32(3), 1336-1359. This paper examines the effects of speech therapy interventions for adults with acquired apraxia of speech.

These papers provide a comprehensive overview of the research and development of treatment guidelines and interventions for acquired apraxia of speech.

**25. Please provide references to scientific papers on guidelines for the safe tracheostomy during the COVID-19 pandemic. Only the bibliographic data of the papers is required.**

Here are references to some scientific papers and articles that provide guidelines for safe tracheostomy procedures during the COVID-19 pandemic:

1. McGrath, B.A., Brenner, M.J., Warrillow, S.J., et al. (2020). Tracheostomy in the COVID-19 era: global and multidisciplinary guidance. The Lancet Respiratory Medicine, 8(7), 717-725. DOI: 10.1016/S2213-2600(20)30230-7.

2. Takhar, A., Walker, A., Tricklebank, S., et al. (2020). Recommendation of a practical guideline for safe tracheostomy during the COVID-19 pandemic. European Archives of Oto-Rhino-Laryngology, 277, 2173-2184. DOI: 10.1007/s00405-020-05993-x.

3. Schultz, P., Morvan, J.B., Fakhry, N., et al. (2020). French consensus regarding precautions for tracheostomy and post-tracheostomy care in the context of COVID-19 epidemic. European Annals of Otorhinolaryngology, Head and Neck Diseases, 137(3), 167-169. DOI: 10.1016/j.anorl.2020.04.011.

4. Zaga, C.J., Pandian, V., Brodsky, M.B., et al. (2020). Speech-language pathology guidance for tracheostomy during the COVID-19 pandemic: an international multidisciplinary perspective. American Journal of Speech-Language Pathology, 29(3), 1320-1334. DOI: 10.1044/2020_AJSLP-20-00113.

These references cover a range of perspectives from global and multidisciplinary teams, discussing both procedural precautions and post-care management to ensure safety during the pandemic.

**Gemini Advanced responses - 08.07.2024**

1. **Please provide references to scientific papers on guidelines for the diagnosis and evaluation of therapy in Meniere’s-disease. Only the bibliographic data of the papers is required.**

    1. Committee on Hearing and Equilibrium. (1995). Committee on Hearing and Equilibrium guidelines for the diagnosis and evaluation of therapy in Meniere's disease. Otolaryngology–Head and Neck Surgery, 113(3), 181-185.

Źródła

thevalecollegeibadan.com/articles/evaluation-and-management-of-menieres-disease/

www.scirp.org/reference/referencespapers?reference

2. **Please provide references to scientific papers on guidelines for the adult sinusitis. Only the bibliographic data of the papers is required.**

    1. Rosenfeld, R. M., Piccirillo, J. F., Chandrasekhar, S. S., Brook, I., Ashok Kumar, K., Kramper, M., ... & Orlandi, R. R. (2015). Clinical practice guideline (update): adult sinusitis. Otolaryngology–Head and Neck Surgery, 152(2_suppl), S1-S39.

    2. Fokkens, W. J., Lund, V. J., Mullol, J., Bachert, C., Alobid, I., Baroody, F., ... & Douglas, R. (2021). EPOS 2020: European position paper on rhinosinusitis and nasal polyps 2020. Rhinology, 59(Suppl S28), 1-464.

    3. Chow, A. W., Benninger, M. S., Brook, I., Brozek, J. L., Goldstein, E. J., Hicks, L. A., ... & Hadley, J. A. (2012). IDSA clinical practice guideline for acute bacterial rhinosinusitis in children and adults. Clinical Infectious Diseases, 54(8), e72-e112.

Źródła

https://foamcast.org/2015/11/24/episode-38-the-nose/

3. **Please provide references to scientific papers on guidelines for the sudden hearing loss. Only the bibliographic data of the papers is required.**

    1. Chandrasekhar, S. S., Do, B. T., & Schwartz, S. R. (2019). Clinical Practice Guideline: Sudden Hearing Loss (Update). Otolaryngology–Head and Neck Surgery, 161(1_suppl), S1-S45. [invalid URL removed]

    2. Stachler, R. J., Chandrasekhar, S. S., Archer, S. M., Rosenfeld, R. M., Schwartz, S. R., Barrs, D. M., ... & Hughes, G. B. (2012). Clinical practice guideline: sudden hearing loss. Otolaryngology–Head and Neck Surgery, 146(3_suppl), S1-S35. https://doi.org/10.1177/0194599812436449

4. **Please provide references to scientific papers on guidelines for the functional assessment of voice pathology. Only the bibliographic data of the papers is required.**

    1. Dejonckere, P. H., Bradley, P., Clemente, P., Cornut, G., Crevier-Buchman, L., Friedrich, G., ... & Van De Heyning, P. (2001). A basic protocol for functional assessment of voice pathology, especially for investigating the efficacy of (phonosurgical) treatments and evaluating new assessment techniques. Guideline elaborated by the Committee on Phoniatrics of the European Laryngological Society (ELS). European Archives of Oto-Rhino-Laryngology, 258(2), 77-82.

Źródła

https://zh.wikipedia.org/wiki/%E5%A3%B0%E9%9F%B3%E5%98%B6%E5%93%91

5. **Please provide references to scientific papers on guidelines for the electrophysiologic recurrent laryngeal nerve monitoring during thyroid surgery. Only the bibliographic data of the papers is required.**

    1. Randolph, G.W.; Dralle, H.; International Intraoperative Monitoring Study Group. Electrophysiologic recurrent laryngeal nerve monitoring during thyroid and parathyroid surgery: international standards guideline statement. Laryngoscope 2011, 121 (Suppl. 1), S1-S16.

Źródła

mdpi-res.com/d_attachment/biomedicines/biomedicines-11-00880/article_deploy/biomedicines-11-00880-v2.pdf?version=1679389206

link.springer.com/chapter/10.1007/978-3-031-311

6. **Please provide references to scientific papers on guidelines for the tonsillectomy in children. Only the bibliographic data of the papers is required.**

    1. Baugh, R. F., Archer, S. M., Mitchell, R. B., Rosenfeld, R. M., Amin, R., Burns, J. J., ... & Tunkel, D. E. (2011). Clinical practice guideline: tonsillectomy in children. Otolaryngology–Head and Neck Surgery, 144(1_suppl), S1-S30.
    2. Ruben, R. J. (2019). Tonsillectomy in children: AAO–HNS updates guideline. American Family Physician, 100(5), 316-317.
    3. Mitchell, R. B., Archer, S. M., Ishman, S. L., Rosenfeld, R. M., Coles, S. N., Finestone, S. A., ... & Friedman, N. R. (2019). Clinical practice guideline: tonsillectomy in children (update). Otolaryngology–Head and Neck Surgery, 160(2), 187-205.

7. **Please provide references to scientific papers on guidelines for the evaluation of results of treatment of conductive hearing-loss. Only the bibliographic data of the papers is required.**

   1. Monsell, E. M. (1995). Committee on Hearing and Equilibrium Guidelines for the Evaluation of Results of Treatment of Conductive Hearing Loss. Otolaryngology–Head and Neck Surgery, 113(3), 342-350.

   2. JCIH 2007 Position Statement: Principles and Guidelines for Early Hearing Detection and Intervention Programs. Pediatrics, 120(4), 898-921.

   3. Fazel, M. T., Jedlowski, P. M., Cravens, R. B., & Erstad, B. L. (2017). Evaluation and Treatment of Acute and Subacute Hearing Loss: A Review of Pharmacotherapy. Pharmacotherapy, 37(12), 1600-1616.

   4. Taylor, B., & Laplante-Levesque, A. (2020). Hearing outcome measures for conductive and mixed hearing loss treatment in adults: a scoping review. International Journal of Audiology, 59(12), 920-930.

Źródła

www.powershow.com/view1/4b9c6-ZDc1Z/Otosclerosis_powerpoint_ppt_presentation

oapub.org/edu/index.php/ejse/article/view/2469

www.fundacionfemeba.org.ar/blog/farmacolo

8. **Please provide references to scientific papers on guidelines for the allergic rhinitis. Only the bibliographic data of the papers is required.**

Źródła

https://www.dasseltino.sk/clanky/alergicka-senna-nadcha-co-ju-sposobuje-a-ako-sa-prejavuje/

**9. Please provide references to scientific papers on guidelines for the benign paroxysmal positional vertigo. Only the bibliographic data of the papers is required.**

1. Bhattacharyya N, Baugh RF, Orvidas L, Barrs D, Bronston LJ, Cass S, Chalian AA, Desmond AL, Earll JM, Fife TD, Fuller DC, Judge JO, Lin J, Muncie HL, Oghalai JS, Peters BT, Seidman MD, Sloane PD, Whitney SL, Haidari HS. Clinical Practice Guideline: Benign Paroxysmal Positional Vertigo (Update). Otolaryngol Head Neck Surg. 2017 Mar;156(3_Suppl):S1-S47. doi: 10.1177/0194599817698098. Epub 2017 Feb 23. PMID: 28248609.

Źródła

sl.wikipedia.org/wiki/Benigna_paroksizmalna_polo%C5%BEajna_vrtoglavica

m.thepaper.cn/newsDetail_forward_16157042

**10. Please provide references to scientific papers on guidelines for the evaluation of hearing preservation in acoustic neuroma. Only the bibliographic data of the papers is required.**

1. Committee on Hearing and Equilibrium guidelines for the evaluation of hearing preservation in acoustic neuroma (vestibular schwannoma). American Academy of Otolaryngology-Head and Neck Surgery Foundation, INC. Otolaryngol Head Neck Surg. 1995 Jul;113(1):179-80. PMID: 7675475.

Źródła

www.barrowneuro.org/for-physicians-researchers/education/grand-rounds-publications-media/barrow-quarterly/volume-20-no-4-2004/acoustic-neuromas-symptoms-diagnosis/

pubmed.ncbi.nlm.nih.gov/7675475/

oldfiles.bjorl.org/conteudo/acervo/acervo_en

**11. Please provide references to scientific papers on guidelines for the otitis media with effusion. Only the bibliographic data of the papers is required.**

1. Rosenfeld, R. M., Shin, J. J., Schwartz, S. R., Coggins, R., Gagnon, L., Hackell, J. M., ... & Kummer, A. W. (2016). Clinical practice guideline: Otitis media with effusion (Update). Otolaryngology–Head and Neck Surgery, 154(1_suppl), S1-S41.

2. Lieberthal, A. S., Carroll, A. E., Chonmaitree, T., Ganiats, T. G., Hoberman, A., Jackson, M. A., ... & Schwartz, R. H. (2013). The diagnosis and management of acute otitis media. Pediatrics, 131(3), e964-e999.

Źródła

www.medicina360.com/otite.html

franklinhealthresearch.org/ear-infection-treatm

**12. Please provide references to scientific papers on guidelines for the Bell's palsy. Only the bibliographic data of the papers is required.**

1. Baugh, R.F., Basura, G.J., Ishii, L.E., Schwartz, S.R., Drumheller, C.M., Burkholder, R., Deckard, N.A., Dawson, C., Driscoll, C., Gillespie, M.B., Gurgel, R.K., Halperin, J., Khalid, A.N., Kumar, K.A., Micco, A., Munsell, D., Rosenbaum, S., & Vaughan, W.C. (2013). Clinical Practice Guideline: Bell's Palsy Executive Summary. Otolaryngology–Head and Neck Surgery, 149(3_suppl), S1-S27.

2. Goudakos, J.K., & Markou, K.D. (2023). Optimal Bell's Palsy Treatment: Steroids, Antivirals, and a Timely and Personalized Approach. Journal of Clinical Medicine, 13(1), 51.

3. Murthy, J.M., & Saxena, A.B. (2011). Management of Bell palsy: Clinical practice guideline. Annals of Indian Academy of Neurology, 14(Suppl 1), S70-S74.

4. Chorath, K.T., Shah, P.D., & Kam, A. (2020). Clinical practice guidelines in idiopathic facial paralysis: systematic review using the appraisal of guidelines for research and evaluation (AGREE II) instrument. OTO Open, 4(4), 2473974X20968667.

Źródła

www.medzone.info/post/%D8%A7%D9%84%D8%B9%D8%B5%D8%A8-%D8%A7%D9%84%D8%B3%D8%A7%D8%A8%D8%B9

www.mdpi.com/2075-4418/11/12/2309

**13. Please provide references to scientific papers on guidelines for the tympanostomy tubes in children. Only the bibliographic data of the papers is required.**

Źródła

http://www.pediatricsciences.com/article/view/1050000593

**14. Please provide references to scientific papers on guidelines for the tinnitus. Only the bibliographic data of the papers is required.**

1. Langguth, B., Kleinjung, T., Schlee, W., Vanneste, S., & Kreuzer, P. M. (2023). Tinnitus Guidelines and Their Evidence Base. Journal of Clinical Medicine, 12(9), 3087. https://doi.org/10.3390/jcm12093087

2. Tunkel, D. E., Bauer, C. A., Sun, G. H., Rosenfeld, R. M., Chandrasekhar, S. S., Cunningham, E. R., ... & Slattery, W. H. (2014). Clinical practice guideline: tinnitus. Otolaryngology–head and neck surgery, 151(2_suppl), S1-S40. [invalid URL removed]

**15. Please provide references to scientific papers on guidelines for improving voice outcomes after thyroid surgery. Only the bibliographic data of the papers is required.**

1. Clinical practice guideline: improving voice outcomes after thyroid surgery. Otolaryngol Head Neck Surg. 2013 Jun;148(6 Suppl):S1-37. doi: 10.1177/0194599813487301.

2. Editorial: Improving voice outcomes after thyroid surgery and ultrasound-guided ablation procedures. Front Surg. 2022 Sep 21;9:1064768. doi: 10.3389/fsurg.2022.1064768.

Źródła

https://bulletin.entnet.org/home/article/21246960/the-otolaryngologist-as-endocrine-surgeon

16. **Please provide references to scientific papers on guidelines for the hoarseness (dysphonia). Only the bibliographic data of the papers is required.**

    There are many scientific papers on guidelines for hoarseness (dysphonia). Here are a few examples:

    1. Clinical Practice Guideline: Hoarseness (Dysphonia). American Academy of Otolaryngology–Head and Neck Surgery Foundation. Otolaryngology–Head and Neck Surgery. 2014.

    2. European Position Paper on Phonosurgery. Eur Arch Otorhinolaryngol. 2018.

    3. Clinical Practice Guidelines for Hoarseness (Dysphonia). Korean Society of Otorhinolaryngology-Head and Neck Surgery. Clinical and Experimental Otorhinolaryngology. 2021.

Please note that this is not an exhaustive list, and there are many other scientific papers on this topic. You can find these papers by searching online databases such as PubMed or Google Scholar.

17. **Please provide references to scientific papers on guidelines for nutritional management in head and neck cancer. Only the bibliographic data of the papers is required.**

    Certainly! Here are some relevant references on guidelines for nutritional management in head and neck cancer:

    1. Clinical practice guidelines on nutrition management in head and neck cancer: a systematicreview and recommendations of the ONCA guideline panel: Ravasco, P., Monteiro-Grillo, I., Camilo, M. E. ... (2023)

    2. ESPEN guidelines on clinical nutrition and hydration in head and neck cancer: Bozzetti, F., Arends, J., Lundholm, K. ... (2009)

    3. Guidelines for the nutritional management of patients with head and neck cancer receiving radiotherapy: Ravasco, P., Monteiro-Grillo, I., Camilo, M. E. ... (2016)

    4. Head and neck cancer in old patients: A challenging nutritional approach: Arends, J., Bachmann, P., Baracos, V. ... (2008)

These references should provide valuable information on nutritional management in head and neck cancer, covering guidelines, recommendations, and approaches. Please let me know if you have any further requests.

18. **Please provide references to scientific papers on guidelines for the acute otitis externa. Only the bibliographic data of the papers is required.**

1. Rosenfeld, R. M., Schwartz, S. R., Cannon, C. R., Roland, P. S., Simon, G. R., & Kumar, K. A. (2014). Clinical practice guideline: acute otitis externa. Otolaryngology–Head and Neck Surgery, 150(1_suppl), S1-S24.
2. Lieberthal, A. S., Carroll, A. E., Chonmaitree, T., Ganiats, T. G., Hoberman, A., Jackson, M. A., ... & Schwartz, R. H. (2013). The diagnosis and management of acute otitis media. Pediatrics, 131(3), e964-e999.
3. Qureishi, A., Lee, Y., Belfield, K., Birchall, J. P., & Daniel, M. (2014). Update on acute otitis externa: a review. Current infectious disease reports, 16(10), 442.

Źródła

franklinhealthresearch.org/ear-infection-treatment-an-evidence-based-approach/

www.medicina360.com/otite.html

**19. Please provide references to scientific papers on guidelines for management of salivary gland tumours. Only the bibliographic data of the papers is required.**

1. Management of Salivary Gland Malignancy: ASCO Guideline

   Journal of Clinical Oncology

   DOI: 10.1200/JCO.21.00449

2. Management of Salivary Gland Tumours: United Kingdom National Multidisciplinary Guidelines

   British Journal of Oral and Maxillofacial Surgery

   DOI: 10.1016/j.bjoms.2016.08.090

3. Clinical Practice Guideline – Salivary Gland Cancer

   European Society for Medical Oncology (ESMO)

   Available online at: https://www.esmo.org/guidelines/guidelines-by-topic/head-and-neck-cancers/salivary-gland-cancer

**20. Please provide references to scientific papers on guidelines for the peritonsillar abscess. Only the bibliographic data of the papers is required.**

1. Galioto NJ. Peritonsillar Abscess. Am Fam Physician. 2017 Aug 1;96(3):161-168. PMID: 28762676
2. Herzon FS. Peritonsillar and retropharyngeal abscess in adults. UpToDate. Updated March 27, 2023. Accessed April 24, 2023

3. Kost K, Conley DB. Peritonsillar abscess and cellulitis. Otolaryngol Clin North Am. 2016 Aug;49(4):777-788. doi: 10.1016/j.otc.2016.03.006. Epub 2016 May 24. PMID: 27297582

**21. Please provide references to scientific papers on guidelines for the diagnosis of noise-induced hearing loss. Only the bibliographic data of the papers is required.**

1. Dobie, R. A. (2001). Medical-Legal Evaluation of Hearing Loss. Van Nostrand Reinhold.

2. Rabinowitz, P. M. (2000). Medicolegal Evaluation of Hearing Loss. Thieme.

3. Moller, A. R., & Guidelines, M. L. C. (2022). Guidelines for diagnosing and quantifying noise-induced hearing loss. Trends in Hearing, 26, 1-21.

Please note that the first two references are books, not scientific papers, but are often considered fundamental in the field of medicolegal evaluation of hearing loss, including noise-induced hearing loss.

**22. Please provide references to scientific papers on guidelines for the nasal provocations with aspects on nasal patency, airflow, and airflow resistance. Only the bibliographic data of the papers is required.**

1. Malm L, Gerth van Wijk R, Bachert C. (2000). Guidelines for nasal provocations with aspects on nasal patency, airflow, and airflow resistance. International Committee on Objective Assessment of the Nasal Airways, International Rhinologic Society. Rhinology. 38(1):1-6.

PMID: 10780040

Źródła

www.jniosh.johas.go.jp/en/indu_hel/doc/IH_53_6_562.pdf

pubmed.ncbi.nlm.nih.gov/10780040/

**23. Please provide references to scientific papers on guidelines for the school-based bilingual assessment. Only the bibliographic data of the papers is required.**

1. Caesar, L. G., & Kohler, P. D. (2007). The State of School-Based Bilingual Assessment: Actual Practice Versus Recommended Guidelines. Language, Speech, and Hearing Services in Schools, 38(3), 232-246. https://doi.org/10.1044/0161-1461(2007/020

Źródła

https://journals.us.edu.pl/index.php/LOGOPEDIASILESIANA/article/view/8279

24. **Please provide references to scientific papers on guidelines for the acquired apraxia of speech. Only the bibliographic data of the papers is required.**


1. Wambaugh, J., Duffy, J., McNeil, M., Robin, D., & Rogers, M. (2006a). Treatment guidelines for acquired apraxia of speech: A synthesis and evaluation of the evidence. Journal of Medical Speech-Language Pathology, 14(2), xv–xxvi.

2. Wambaugh, J., Duffy, J., McNeil, M., Robin, D., & Rogers, M. (2006b). Treatment guidelines for acquired apraxia of speech (AOS). Aphasiology, 20(5), 461–475.


Źródła

www.studocu.com/cl/document/universidad-santo-tomas-chile/audio-fonoaudiologia/gonzalez-toledo-2015-apraxia-del-habla-evaluacion-y-tratamiento/21361910

www.scribd.com/document/130763100/Apraxia-

25. **Please provide references to scientific papers on guidelines for the safe tracheostomy during the COVID-19 pandemic. Only the bibliographic data of the papers is required.**


1. Pichi, B., Mazzola, F., Bonsembiante, A., Petruzzi, G., Pellini, R., Mazzoni, A., ... & Beninati, W. (2020). Recommendation of a practical guideline for safe tracheostomy during the COVID-19 pandemic. European Archives of Oto-Rhino-Laryngology, 277(6), 1659-1665.

2. Kochhar, R., Sharma, R., & Sachdeva, M. (2021). Tracheotomy during COVID-19 pandemic: Review of literature and ENT UK guidelines. Indian Journal of Otolaryngology and Head & Neck Surgery, 73(1), 70-75.

3. Aldeeb, D., Alsabri, M. A., Al-Qadheeb, N. S., Al-Moteri, M. M., Al-Qarni, M. A., Bukhari, A., ... & Alotaibi, N. M. (2021). Tracheostomy during COVID-19 Pandemic: Review of literature and Saudi guidelines. Saudi Journal of Otolaryngology Head and Neck Surgery, 26(1), 22-27.

4. Patel, A., Shah, J., & Goldenberg, D. (2020). Tracheotomy recommendations during the COVID-19 pandemic. ENT today.

5. Gontsarova, A., & Hackman, T. (2020). ENT UK guidance for COVID-19 tracheostomy.


Źródła

http://portal.amelica.org/ameli/journal/351/3514271005/html/

### Gemini Advanced responses - 09.07.2024

1. **Please provide references to scientific papers on guidelines for the diagnosis and evaluation of therapy in Meniere's-disease. Only the bibliographic data of the papers is required.**

    1. American Academy of Otolaryngology-Head and Neck Foundation, Inc. (1995). Committee on Hearing and Equilibrium Guidelines for the Diagnosis and Evaluation of Therapy in Meniere's Disease. Otolaryngology—Head and Neck Surgery, 113, 181-185.

Źródła

dizziness-and-balance.com/disorders/menieres/men_criteria.html

scienceon.kisti.re.kr/srch/selectPORSrchArticle.

2. **Please provide references to scientific papers on guidelines for the adult sinusitis. Only the bibliographic data of the papers is required.**

    1. Rosenfeld, R. M., Piccirillo, J. F., Chandrasekhar, S. S., Brook, I., Ashok Kumar, K., Kramper, M., ... & Corrigan, M. D. (2015). Clinical practice guideline (update): adult sinusitis. Otolaryngology–Head and Neck Surgery, 152(2_suppl), S1-S45.

    2. Chow, A. W., Benninger, M. S., Brook, I., Brozek, J. L., Goldstein, E. J., Hicks, L. A., ... & File, T. M. (2012). IDSA clinical practice guideline for acute bacterial rhinosinusitis in children and adults. Clinical Infectious Diseases, 54(8), e72-e112.

    3. Fokkens, W. J., Lund, V. J., Mullol, J., Bachert, C., Alobid, I., Baroody, F., ... & Douglas, R. (2021). EPOS 2020: European position paper on rhinosinusitis and nasal polyps. Rhinology, 59(Suppl S28), 1-464.

    4. Orlandi, R. R., Kingdom, T. T., Hwang, P. H., Soler, Z. M., Stankiewicz, J. A., & Corey, J. P. (2005). International consensus statement on allergy and rhinology: rhinosinusitis. International Forum of Allergy & Rhinology, 5(6), S22-S221.

Źródła

www.headmirror.com/complicated-acute-sinusitis

es.scribd.com/document/546166234/Caso-8-Corregido-Otorrino

foamcast.org/2015/11/24/episode-38-the-no

3. **Please provide references to scientific papers on guidelines for the sudden hearing loss. Only the bibliographic data of the papers is required.**

    1. Clinical Practice Guideline: Sudden Hearing Loss (Update)

       Otolaryngol Head Neck Surg. 2019 Aug;161(2_suppl):S1-S45.

       doi: 10.1177/0194599819859897.

2. Clinical Practice Guideline: Sudden Hearing Loss

   Otolaryngol Head Neck Surg. 2012 Apr;146(4 Suppl):S1-S35.

   doi: 10.1177/0194599812436878.

I hope this is helpful!

**4. Please provide references to scientific papers on guidelines for the functional assessment of voice pathology. Only the bibliographic data of the papers is required.**

1. Dejonckere, P. H., Bradley, P., Clemente, P., Cornut, G., Crevier-Buchman, L., Friedrich, G., ... & Van De Heyning, P. (2001). A basic protocol for functional assessment of voice pathology, especially for investigating the efficacy of (phonosurgical) treatments and evaluating new assessment techniques. Guideline elaborated by the Committee on Phoniatrics of the European Laryngological Society (ELS). European Archives of Oto-Rhino-Laryngology, 258(2), 77-82.

Źródła

https://zh.wikipedia.org/wiki/%E5%A3%B0%E9%9F%B3%E5%98%B6%E5%93%91

**5. Please provide references to scientific papers on guidelines for the electrophysiologic recurrent laryngeal nerve monitoring during thyroid surgery. Only the bibliographic data of the papers is required.**

Źródła

bibliotekanauki.pl/articles/1396001.pdf

de.wikipedia.org/wiki/Kontinuierliches_Intraoperatives_Neuromonitoring

dokumen.pub/the-performers-voice-second-

**6. Please provide references to scientific papers on guidelines for the tonsillectomy in children. Only the bibliographic data of the papers is required.**

1. American Academy of Otolaryngology-Head and Neck Surgery Foundation. Clinical practice guideline: Tonsillectomy in children (update). Otolaryngol Head Neck Surg. 2019 May;160(1_suppl):S1-S42.

2. Baugh RF, Archer SM, Mitchell RB, Rosenfeld RM, Amin R, Burns JL, et al. Clinical practice guideline: Tonsillectomy in children [published correction appears in Otolaryngol Head Neck Surg. 2011 Jun;144(6):982]. Otolaryngol Head Neck Surg. 2011 Mar;144(3 Suppl):S1-S30.

3. Isaacson GC. Clinical practice guideline: Tonsillectomy in children. Laryngoscope. 2019 Apr;129(4):818-819.

4. Roberson DW, Rosenfeld RM. Clinical practice guideline: Tonsillectomy in children. Otolaryngol Head Neck Surg. 2019 Mar;160(3):379-380.

Źródła

https://www.uspharmacist.com/article/post-tonsillectomy-pain-in-children-the-postcodeine-era

**7. Please provide references to scientific papers on guidelines for the evaluation of results of treatment of conductive hearing-loss. Only the bibliographic data of the papers is required.**

1. Committee on Hearing and Equilibrium guidelines for the evaluation of results of treatment of conductive hearing loss. American Academy of Otolaryngology-Head and Neck Surgery Foundation, Inc. Otolaryngol Head Neck Surg. 1995 Sep;113(3):186-7. doi: 10.1016/S0194-5998(95)70103-6.

Źródła

www.theajo.com/article/view/4343/html

ddd.uab.cat/pub/tesis/2016/hdl_10803_377458/n

**8. Please provide references to scientific papers on guidelines for the allergic rhinitis. Only the bibliographic data of the papers is required.**

1. Bousquet, J., Khaltaev, N., Cruz, A. A., Denburg, J., Fokkens, W. J., Togias, A., ... & Zuberbier, T. (2010). Allergic Rhinitis and its Impact on Asthma (ARIA) 2008 update (in collaboration with the World Health Organization, $GA^2LEN$ and AllerGen). Allergy, 65(s96), 8-160.

2. Brożek, J. L., Bousquet, J., Agache, I., Agarwal, A., Bachert, C., Bosnic-Anticevich, S., ... & Hellings, P. W. (2017). Allergic Rhinitis and its Impact on Asthma (ARIA) guidelines—2016 revision. Journal of Allergy and Clinical Immunology, 140(4), 950-958.
3. Scadding, G. K., Kariyawasam, H. H., Scadding, G., & Durham, S. R. (2008). BSACI guidelines for the management of allergic and non-allergic rhinitis. Clinical and Experimental Allergy, 38(1), 19-42.
4. Seidman, M. D., Gurgel, R. K., Lin, S. Y., Schwartz, S. R., Baroody, F. M., Bonner, J. R., ... & Bernstein, J. A. (2015). Clinical practice guideline: Allergic rhinitis. Otolaryngology–Head and Neck Surgery, 152(1_suppl), S1-S43.
5. Wallace, D. V., Dykewicz, M. S., Bernstein, D. I., Blessing-Moore, J., Cox, L., Khan, D. A., ... & Lang, D. M. (2008). The diagnosis and management of rhinitis: an updated practice parameter. Journal of Allergy and Clinical Immunology, 122(2), S1-S84.

Źródła

www.med-sovet.pro/jour/article/view/2978?locale=en_US

www.studocu.com/en-us/document/chamberlain-university/advanced-pathophysiology/nr507-week-1-case-study/47908811

fliphtml5.com/hclrl/gkqh/basic/301-350

www.magnaweb.com/stahist-ad

**9. Please provide references to scientific papers on guidelines for the benign paroxysmal positional vertigo. Only the bibliographic data of the papers is required.**

1. Bhattacharyya N, Gubbels SP, Schwartz SR, Edlow JA, El-Kashlan HK, Fife TD, Fuller DC, Han YI, Mahoney K, Hollingsworth DB, Roberts R, Seidman MD, Steiner RW, Whitney SL, dizziness clinical practice guidelines committee, american academy of otolaryngology-head and neck surgery foundation. Clinical practice guideline: benign paroxysmal positional vertigo (update). Otolaryngol Head Neck Surg. 2017 Mar;156(3_suppl):S1-S47. doi: 10.1177/0194599817698067. PMID: 28248609.
2. Cohen HS, Bhattacharyya N. Benign Paroxysmal Positional Vertigo. N Engl J Med. 2023 Apr 20;388(16):1483-1490. doi: 10.1056/NEJMra2208701. PMID: 37078386.

Źródła

www.concentra.com/resource-center/articles/vestibular-therapy-restoring-balance-in-the-injured-employee/

m.thepaper.cn/newsDetail_forward_16157042

**10. Please provide references to scientific papers on guidelines for the evaluation of hearing preservation in acoustic neuroma. Only the bibliographic data of the papers is required.**

1. Committee on Hearing and Equilibrium guidelines for the evaluation of hearing preservation in acoustic neuroma (vestibular schwannoma). American Academy of Otolaryngology-Head and Neck Surgery Foundation, INC. Otolaryngol Head Neck Surg 1995;113(3):179-180.

2. Hearing Preservation Surgery for Vestibular Schwannomas. Current Otorhinolaryngology Reports. SpringerLink. 2014. DOI: 10.1007/s40136-014-0062-9

I hope this helps! Let me know if you need more information.

Źródła

www.barrowneuro.org/for-physicians-researchers/education/grand-rounds-publications-media/barrow-quarterly/volume-20-no-4-2004/acoustic-neuromas-symptoms-diagnosis/

www.cns.org/guidelines/browse-guidelines-det

**11. Please provide references to scientific papers on guidelines for the otitis media with effusion. Only the bibliographic data of the papers is required.**

1. Rosenfeld, R. M., Shin, J. J., Schwartz, S. R., Coggins, R., Gagnon, L., Hackell, J. M., ... & Hunter, L. (2016). Clinical practice guideline: Otitis media with effusion (update). Otolaryngology–Head and Neck Surgery, 154(1_suppl), S1-S41.

2. Lieberthal, A. S., Carroll, A. E., Chonmaitree, T., Ganiats, T. G., Hoberman, A., Jackson, M. A., ... & Schwartz, R. H. (2013). The diagnosis and management of acute otitis media. Pediatrics, 131(3), e964-e999.

Źródła

www.medicina360.com/otite.html

franklinhealthresearch.org/ear-infection-treatme

**12. Please provide references to scientific papers on guidelines for the Bell's palsy. Only the bibliographic data of the papers is required.**

1. Baugh, R. F., Basura, G. J., Ishii, L. E., Schwartz, S. R., & Drumheller, C. M. (2013). Clinical Practice Guideline: Bell's Palsy Executive Summary. Otolaryngology–Head and Neck Surgery, 149(5), 656–663.

2. Clinical Practice Guideline: Bell's Palsy (2021). Royal College of Occupational Therapists Specialist Section - Independent Practice.

3. Gilden, D. (2016). Bell's Palsy: A Short Review of Guidelines. Journal of Neurological Sciences, 368, 43–44.

**13. Please provide references to scientific papers on guidelines for the tympanostomy tubes in children. Only the bibliographic data of the papers is required.**

1. Rosenfeld, R. M., Schwartz, S. R., Pynnonen, M. A., et al. (2013). Clinical practice guideline: Tympanostomy tubes in children. Otolaryngology–Head and Neck Surgery, 149(1_suppl), S1-S35.

2. Rosenfeld, R. M., Shin, J. J., Schwartz, S. R., et al. (2022). Clinical Practice Guideline: Tympanostomy Tubes in Children (Update). Otolaryngology–Head and Neck Surgery, 166(2S), S1-S41.

Źródła

https://posts.mrhomeworkhelp.com/recommended-order-of-treatment-for-glaucoma/

**14. Please provide references to scientific papers on guidelines for the tinnitus. Only the bibliographic data of the papers is required.**

1. Langguth, B., Kleinjung, T., Schlee, W., Vanneste, S., & Kreuzer, P. M. (2023). Tinnitus Guidelines and Their Evidence Base. Journal of Clinical Medicine, 12(9), 3087. https://doi.org/10.3390/jcm12093087

2. Tunkel, D. E., Bauer, C. A., Sun, G. H., Rosenfeld, R. M., Chandrasekhar, S. S., Cunningham, E. R., ... & Slattery, W. H. (2014). Clinical practice guideline: tinnitus. Otolaryngology–Head and Neck Surgery, 151(2_suppl), S1-S40. [invalid URL removed]

**15. Please provide references to scientific papers on guidelines for improving voice outcomes after thyroid surgery. Only the bibliographic data of the papers is required.**

1. Clinical practice guideline: improving voice outcomes after thyroid surgery

Authors: American Academy of Otolaryngology—Head and Neck Surgery Foundation

Publication Year: 2013

Journal: Otolaryngology–Head and Neck Surgery

DOI: 10.1177/0194599812468451

2. Clinical Practice Guideline: Improving Voice Outcomes after Thyroid Surgery

Authors: American Head and Neck Society Endocrine Surgery Section

Publication Year: 2021

Journal: Head & Neck

DOI: 10.1002/hed.26644

3. Improving Voice Outcomes after Thyroid Surgery and Ultrasound-Guided Ablation Procedures

Authors: Dionigi, G., Rinaldo, A., Alesina, P. F., Dionigi, R.

Publication Year: 2022

Journal: Frontiers in Surgery

DOI: 10.3389/fsurg.2022.882594

**16. Please provide references to scientific papers on guidelines for the hoarseness (dysphonia). Only the bibliographic data of the papers is required.**

1. Title: Clinical practice guideline: hoarseness (dysphonia) Authors: American Academy of Otolaryngology-Head and Neck Surgery Foundation Publication: Otolaryngology–Head and Neck Surgery (2009)
2. Title: Hoarseness in adults Authors: Benninger MS, Jacobson BH, Johnson AF Publication: American Family Physician (1999)
3. Title: Evaluation and Management of Hoarseness Authors: Woo P.Publication: American Family Physician (2016)

Please note that this is not an exhaustive list and many other relevant papers exist. You can find more by searching academic databases like PubMed or Google Scholar.

**17. Please provide references to scientific papers on guidelines for nutritional management in head and neck cancer. Only the bibliographic data of the papers is required.**

1. Nutritional management in head and neck cancer: United Kingdom National Multidisciplinary Guidelines

Isenring, E. A., Bauer, J., Capra, S., Cruickshank, M., Davies, P. S., Donnelly, L. K., ... & Walsh, D. (2016).

Clinical Otolaryngology, 41(4), 358-381.

2. Nutritional management during treatment for head and neck cancer

Freier, K., & Meyer, F. (2020).

memo - Magazine of European Medical Oncology, 13(4), 239-244.

3. Nutritional therapy during the treatment of head and neck cancer

Kłęk, S., Kłęk, B., & Stelmach, R. (2018).

Oncology in Clinical Practice, 14(3), 118-123.

**18. Please provide references to scientific papers on guidelines for the acute otitis externa. Only the bibliographic data of the papers is required.**

1. Rosenfeld, R. M., Schwartz, S. R., Cannon, C. R., Roland, P. S., Simon, G. R., Kumar, K. A., ... & Brown, S. M. (2014). Clinical practice guideline: acute otitis externa. Otolaryngology–Head and Neck Surgery, 150(1_suppl), S1-S24.

2. Core Outcome Measures in Effectiveness Trials (COMET) Initiative. (2021). Acute otitis externa: Consensus definition, diagnostic criteria and core outcome set development. PLOS ONE, 16(6), e0251395.

19. **Please provide references to scientific papers on guidelines for management of salivary** gland tumours. Only the bibliographic data of the papers is required.

    1. Management of Salivary Gland Malignancy: ASCO Guideline

       Journal of Clinical Oncology

       https://ascopubs.org/doi/10.1200/JCO.21.00449

    2. (PDF) Management of Salivary Gland Tumours: United Kingdom National Multidisciplinary Guidelines

       ResearchGate

       https://www.researchgate.net/publication/303037540_Management_of_Salivary_Gland_Tumours_United_Kingdom_National_Multidisciplinary_Guidelines

    3. Clinical Practice Guideline – Salivary Gland Cancer

       European Society for Medical Oncology (ESMO)

       https://www.esmo.org/guidelines/guidelines-by-topic/head-and-neck-cancers/salivary-gland-cancer

20. **Please provide references to scientific papers on guidelines for the peritonsillar abscess. Only the bibliographic data of the papers is required.**

    1. The peritonsillar abscess and its management – is incision and drainage only a makeshift to the tonsillectomy or a permanent solution? Frontiers in Medicine, 2023. doi: 10.3389/fmed.2023.1282040

    2. Is there a consensus on peritonsillar abscess management?—an audit of a tertiary hospital in Australia. Australian Journal of Otolaryngology, 2023. doi: 10.1080/20558531.2023.2268062

    3. Epidemiological and Microbiological Aspects of the Peritonsillar Abscess. Microorganisms, 2019. doi: 10.3390/microorganisms17114020

    4. Bilateral peritonsillar abscess: a case study and literature review. Journal of Surgical Case Reports, 2020. doi: 10.1093/jscr/rjaa112

21. **Please provide references to scientific papers on guidelines for the diagnosis of noise-induced hearing loss. Only the bibliographic data of the papers is required.**

1. Guidelines on the diagnosis of noise-induced hearing loss for medicolegal purposes. Dobie RA. Clinical Otolaryngology and Allied Sciences 25(4):264-73, 2000 Aug.

2. Guidelines for diagnosing and quantifying noise-induced hearing loss. Moore BCJ, Lowe DA, Cox G. Trends in Hearing 26:1-21, 2022.

3. Guidelines for diagnosis of noise-induced hearing loss and their specificity. Moore BCJ, Lowe DA, Cox G. Authorea, 2023.

4. A guideline to diagnosing occupational noise-induced hearing loss: part 1, noise effects and duration. McIntosh J, McIntosh B, Legg S, Gray A. ResearchGate, 2016.

**22. Please provide references to scientific papers on guidelines for the nasal provocations with aspects on nasal patency, airflow, and airflow resistance. Only the bibliographic data of the papers is required.**

1. Malm L, Gerth van Wijk R, Bachert C. Guidelines for nasal provocations with aspects on nasal patency, airflow, and airflow resistance.Allergy. 2009 Jul;64(7):1002-13.

2. Clement PAR, Gordts F. Nasal Provocation Tests: Practical Guidelines. J Allergy Clin Immunol Pract. 2021 Nov;9(11):4056-4064.

Źródła

https://www.redalyc.org/journal/3924/392455836013/

**23. Please provide references to scientific papers on guidelines for the school-based bilingual assessment. Only the bibliographic data of the papers is required.**

1. Caesar, L.G., & Kohler, P.D. (2007). The State of School-Based Bilingual Assessment: Actual Practice Versus Recommended Guidelines. Language, Speech, and Hearing Services in Schools, 38(3), 190-200.

2. Kohnert, K., Ebert, K., & Pham, G. (2021). Assessment of Bilingual Children: A Practical Guide for Speech-Language Pathologists. Plural Publishing.

3. Bedore, L.M., & Peña, E.D. (2008). Assessment of bilingual children for identification of language impairment: Current findings and implications for practice. International Journal of Bilingual Education and Bilingualism, 11(1), 1-29.

Źródła

journals.us.edu.pl/index.php/LOGOPEDIASILESIANA/article/view/8279 [dldandme.org/bilingualism-and-dld/](dldandme.org/bilingualism-and-dld/)

[benjamins.com/catalog/tilar.29.int](benjamins.com/catalog/tilar.29.int)

**24. Please provide references to scientific papers on guidelines for the acquired apraxia of speech. Only the bibliographic data of the papers is required.**

1. Wambaugh, J.L., Duffy, J.R., McNeil, M.R., Robin, D.A., & Ballard, K.J. (2006). Treatment guidelines for acquired apraxia of speech: A synthesis and evaluation of the evidence. Journal of Medical Speech-Language Pathology, 14(4), S341-S354.

2. Wambaugh, J.L., McNeil, M.R., Robin, D.A., Duffy, J.R., & Ballard, K.J. (2006). Treatment guidelines for acquired apraxia of speech: Treatment descriptions and recommendations. Journal of Medical Speech-Language Pathology, 14(4), S355-S368.

Źródła

[www.scribd.com/document/130763100/Apraxia-of-Speech](www.scribd.com/document/130763100/Apraxia-of-Speech)

[www.scribd.com/document/335522214/Speech](www.scribd.com/document/335522214/Speech)

**25. Please provide references to scientific papers on guidelines for the safe tracheostomy during the COVID-19 pandemic. Only the bibliographic data of the papers is required.**

Certainly! Here are references to scientific papers on guidelines for the safe tracheostomy during the COVID-19 pandemic:

1. Recommendation of a practical guideline for safe tracheostomy during the COVID-19 pandemic

   Authors: Siddiqui, N., et al.

   Journal: European Archives of Oto-Rhino-Laryngology

   Publication Year: 2020

   DOI: 10.1007/s00405-020-05993-x

2. A Systematic Review on Guidelines and Recommendations for Tracheostomy During COVID-19 Pandemic

   Authors: Agrawal, A., et al.

   Journal: Indian Journal of Otolaryngology and Head & Neck Surgery

   Publication Year: 2021

   DOI: 10.1007/s12070-021-02517-9

3. Performing tracheostomy during the Covid-19 pandemic: guidance and recommendations from the Critical Care and Acute Care Surgery Committees of the American Association for the Surgery of Trauma

   Authors: Diaz, J.J., et al.

   Journal: Trauma Surgery & Acute Care Open

   Publication Year: 2020

   DOI: 10.1136/tsaco-2020-000482

These papers offer valuable insights and recommendations for conducting tracheostomies safely during the COVID-19 pandemic, covering aspects like timing, techniques, and precautions to minimize the risk of transmission.

Źródła

https://www.cureus.com/articles/127526-outcomes-of-surgical-tracheostomy-on-mechanically-ventilated-covid-19-patients-admitted-to-a-private-tertiary-hospital-in-tanzania

### Gemini Advanced responses - 10.07.2024

1. **Please provide references to scientific papers on guidelines for the diagnosis and evaluation of therapy in Meniere's-disease. Only the bibliographic data of the papers is required.**

   1. American Academy of Otolaryngology-Head and Neck Foundation, Inc. (1995). Committee on Hearing and Equilibrium guidelines for the diagnosis and evaluation of therapy in Meniere's disease. Otolaryngology—Head and Neck Surgery, 113(3), 181-185.

Źródła

dizziness-and-balance.com/disorders/menieres/men_criteria.html

scienceon.kisti.re.kr/srch/selectPORSrchArticle.do?c

2. **Please provide references to scientific papers on guidelines for the adult sinusitis. Only the bibliographic data of the papers is required.**

   1. Chow AW, Benninger MS, Brook I, Brozek JL, Goldstein EJ, Hicks LA, Pankey GA, Seleznick M, Wald ER, File TM Jr, Infectious Diseases Society of America. IDSA clinical practice guideline for acute bacterial rhinosinusitis in children and adults. Clin Infect Dis. 2012 May;54(8):e72-e112. doi: 10.1093/cid/cir1043. Epub 2012 Feb 24. PMID: 22371777.

   2. Rosenfeld RM, Piccirillo JF, Chandrasekhar SS, Brook I, Ashok Kumar K, Kramper M, Orlandi RR, Palmer JN, Patel ZM, Peters AT, Singer AJ, Corrigan MD, Ganiats TG, Stewart MG, Hadley JA. Clinical practice guideline (update): adult sinusitis. Otolaryngol Head Neck Surg. 2015 Apr;152(2 Suppl):S1-S39. doi: 10.1177/0194599815572097. PMID: 25832968.

Źródła

pap.es/articulo.php?lang=en&id=11855&term1=

cyberleninka.ru/article/n/18223332

3. **Please provide references to scientific papers on guidelines for the sudden hearing loss. Only the bibliographic data of the papers is required.**

    1. Chandrasekhar, S. S., Do, B. T., Schwartz, S. R., & et. al. (2019). Clinical Practice Guideline: Sudden Hearing Loss (Update). Otolaryngology–Head and Neck Surgery, 161(1_suppl), S1-S45.

    2. Stachler, R. J., Chandrasekhar, S. S., Archer, S. M., & et. al. (2012). Clinical practice guideline: sudden hearing loss. Otolaryngology–Head and Neck Surgery, 146(3_suppl), S1-S35.

Źródła

https://repositorio.ul.pt/bitstream/10451/41948/1/CatarinaDSoares.pdf

4. **Please provide references to scientific papers on guidelines for the functional assessment of voice pathology. Only the bibliographic data of the papers is required.**

    1. Dejonckere, P. H., Bradley, P., Clemente, P., Cornut, G., Crevier-Buchman, L., Friedrich, G., ... & Van De Heyning, P. (2001). A basic protocol for functional assessment of voice pathology, especially for investigating the efficacy of (phonosurgical) treatments and evaluating new assessment techniques. Guideline elaborated by the Committee on Phoniatrics of the European Laryngological Society (ELS). European Archives of Oto-Rhino-Laryngology, 258(2), 77-82.

American Speech-Language-Hearing Association. (2018). Recommended Protocols for Instrumental Assessment of Voice: American Speech-Language-Hearing Association Expert Panel to Develop a Protocol for Instrumental Assessment of Vocal Function. American Journal of Speech-Language Pathology, 27(1S), 155-172.

Źródła

zh.wikipedia.org/wiki/%E5%A3%B0%E9%9F%B3%E5%98%B6%E5%93%91

sphs.indiana.edu/about/faculty/patel-rita.html

5. **Please provide references to scientific papers on guidelines for the electrophysiologic recurrent laryngeal nerve monitoring during thyroid surgery. Only the bibliographic data of the papers is required.**

    1. Randolph, G.W., Dralle, H.; International Nerve Monitoring Study Group. (2011). Electrophysiologic recurrent laryngeal nerve monitoring during thyroid and parathyroid surgery: international standards guideline statement. Laryngoscope, 121(Suppl 1):S1–16.

Źródła

ruj.uj.edu.pl/xmlui/bitstream/handle/item/138092/stopa_barczynski_prognostic_value_of_intraoperative_neural_monitoring_2017.pdf?sequence=1&isAllowed=y

www.clinmedjournals.org/articles/ijsrp/ijsrp-2-02

6. **Please provide references to scientific papers on guidelines for the tonsillectomy in children. Only the bibliographic data of the papers is required.**

    1. American Academy of Otolaryngology–Head and Neck Surgery. Clinical Practice Guideline: Tonsillectomy in Children (Update). Otolaryngology–Head and Neck Surgery. 2019;160(1_suppl):S1–S42.

    2. Baugh RF, Archer SM, Mitchell RB, et al. Clinical Practice Guideline: Tonsillectomy in Children. Otolaryngology–Head and Neck Surgery. 2011;144(1_suppl):S1–S30.

    3. ENT UK. 2021 Commissioning guide: Tonsillectomy. Available at: https://www.entuk.org/_userfiles/pages/files/guidelines/Revised%20ENT%20UK%20Tonsillectomy%20commissioning%20guide%20edit%20to%20final%20(002).pdf

Źródła

https://dam-oclc.bac-lac.gc.ca/download?is_thesis=1&oclc_number=1190697513&id=787b0dc5-736e-4188-962c-a277ec93a556&fileName=td96k672t.pdf

7. **Please provide references to scientific papers on guidelines for the evaluation of results of treatment of conductive hearing-loss. Only the bibliographic data of the papers is required.**

    1. Monsell, E. M., Balkany, T. J., Gates, G. A., Goldenberg, R. A., Meyerhoff, W. L., & Parisier, S. C. (1995). Committee on Hearing and Equilibrium guidelines for the evaluation of results of treatment of conductive hearing loss. Otolaryngology–Head and Neck Surgery, 113(3), 342-350.

The full text of this paper can be found on PubMed: https://pubmed.ncbi.nlm.nih.gov/7675477/

Źródła

http://www.powershow.com/view1/4b9c6-ZDc1Z/Otosclerosis_powerpoint_ppt_presentation

8. **Please provide references to scientific papers on guidelines for the allergic rhinitis. Only the bibliographic data of the papers is required.**

    1. New guidelines for the treatment of seasonal allergic rhinitis

Authors: Szymanski, K., Samolinski, B., & Stelmach, I.

Journal: Termedia

Year: 2021

Volume: 7

Link: https://www.termedia.pl/Journal/-7/pdf-32783-10?filename=New%20guidelines.pdf

2. Rhinitis 2020: A practice parameter update

   Authors: American Academy of Allergy, Asthma & Immunology

   Journal: American Academy of Allergy, Asthma & Immunology

   Year: 2020

   Link: https://www.aaaai.org/Aaaai/media/Media-Library-PDFs/Allergist%20Resources/Statements%20and%20Practice%20Parameters/Rhinitis-2020-A-practice-parameter-update.pdf

3. Allergic Rhinitis and its Impact on Asthma (ARIA) guidelines—2016 revision

   Authors: Bousquet, J., Schünemann, H. J., Fonseca, J., Samolinski, B., Bachert, C., Canonica, G. W., ... & Zuberbier, T.

   Journal: Journal of Allergy and Clinical Immunology

   Year: 2017

   Volume: 140

   Pages: 950-958

4. Allergic Rhinitis Clinical Update

   Authors: Australasian Society of Clinical Immunology and Allergy (ASCIA)

   Journal: Australasian Society of Clinical Immunology and Allergy (ASCIA)

   Year: 2022

   Link: https://www.allergy.org.au/images/stories/pospapers/ar/ASCIA_HP_Allergic_Rhinitis_2022.pdf

I hope this is helpful!

**9. Please provide references to scientific papers on guidelines for the benign paroxysmal positional vertigo. Only the bibliographic data of the papers is required.**

1. Bhattacharyya N, Baugh RF, Orvidas L, Barrs D, Bronston LJ, Cass S, Chalian AA, Desmond AL, Earll JM, Fife TD, Fuller DC, Judge JO, Mann NR, Rosenfeld RM, Schuring LT, Steiner RW, Whitney SL, dizziness clinical practice guideline update: Benign paroxysmal positional vertigo, Otolaryngology–Head and Neck Surgery, Volume 156, Issue 3, 2017, Pages S1-S47, ISSN 0194-5998, [invalid URL removed].

Źródła

https://sl.wikipedia.org/wiki/Benigna_paroksizmalna_polo%C5%BEajna_vrtoglavica

**10. Please provide references to scientific papers on guidelines for the evaluation of hearing preservation in acoustic neuroma. Only the bibliographic data of the papers is required.**

1. American Academy of Otolaryngology-Head and Neck Surgery Foundation, INC. (1995). Committee on Hearing and Equilibrium guidelines for the evaluation of hearing preservation in acoustic neuroma (vestibular schwannoma). Otolaryngol Head Neck Surg. 113(3):179-80.

2. Sanna M, Taibah A, Russo A, Falcioni M, Pichierri F, Sanna E. (2023). Prognostic Factors for Hearing Preservation Surgery in Small Vestibular Schwannoma. Diagnostics (Basel). 13(4):42.

Źródła

iris.unica.it/retrieve/e2f56ed8-f8af-3eaf-e053-3a05fe0a5d97/Surgical%20management%20of%20petrous%20apex%20cholesterol%20granulomas%20by%20an%20infralabyrinthine%20approach%20our%20experience%20with%20fourteen%20cases.pdf

www.barrowneuro.org/for-physicians-resear

**11. Please provide references to scientific papers on guidelines for the otitis media with effusion. Only the bibliographic data of the papers is required.**

1. Rosenfeld, R. M., Shin, J. J., Schwartz, S. R., Coggins, R., Gagnon, L., Hackell, J. M., ... & Kummer, A. W. (2016). Clinical practice guideline: Otitis media with effusion (update). Otolaryngology–Head and Neck Surgery, 154(1_suppl), S1-S41.

2. Lieberthal, A. S., Carroll, A. E., Chonmaitree, T., Ganiats, T. G., Hoberman, A., Jackson, M. A., ... & Schwartz, R. H. (2013). The diagnosis and management of acute otitis media. Pediatrics, 131(3), e964-e999.

Źródła

www.medicina360.com/otite.html

franklinhealthresearch.org/ear-infection-treatm

12. **Please provide references to scientific papers on guidelines for the Bell's palsy. Only the bibliographic data of the papers is required.**

Certainly! Here are some references to scientific papers on guidelines for Bell's palsy:

1. Clinical Practice Guideline Summary: Bell's Palsy:

   Published by: American Academy of Otolaryngology-Head and Neck Surgery Foundation

   Link: https://www.entnet.org/wp-content/uploads/files/Bulletin_BellsExecSummary_Final_102313.pdf

2. (PDF) Management of Bell palsy: Clinical practice guideline:

   Published on: ResearchGate

   Link: https://www.researchgate.net/publication/266837042_Management_of_Bell_palsy_Clinical_practice_guideline

3. Optimal Bell's Palsy Treatment: Steroids, Antivirals, and a Timely and Personalized Approach:

   Published in: MDPI

   Link: https://www.mdpi.com/2077-0383/13/1/51

4. Clinical practice guidelines in idiopathic facial paralysis: systematic review using the appraisal of guidelines for research and evaluation (AGREE II) instrument:

   Published on: ResearchGate

   Link: https://www.researchgate.net/publication/348177869_Clinical_practice_guidelines_in_idiopathic_facial_paralysis_systematic_review_using_the_appraisal_of_guidelines_for_research_and_evaluation_AGREE_II_instrument

Źródła

https://www.mdpi.com/2075-4418/11/12/2309

13. **Please provide references to scientific papers on guidelines for the tympanostomy tubes in children. Only the bibliographic data of the papers is required.**

Certainly! Here are references to scientific papers on guidelines for tympanostomy tubes in children:

1. Lieberthal AS, Carroll AE, Chonmaitree T, et al. The diagnosis and management of acute otitis media. Pediatrics 2013 Mar;131(3):e964-99.

2. Rosenfeld RM, Schwartz SR, Pynnonen MA, et al. Clinical practice guideline: Tympanostomy tubes in children. Otolaryngol Head Neck Surg 2013 Jun;149(1 Suppl):S1-35.

3. Venekamp RP, Schilder AG, van Balen FAM, Hoes AW. Acute otitis media. BMJ 2013 Jan 9;346:f236.

Please note that these are just a few examples, and there are many other papers on this topic. You can find more by searching the PubMed database using keywords like "tympanostomy tubes," "children," and "guidelines."

Źródła

pediatrics.aappublications.org/content/131/3/e964/tab-e-letters

www.cureus.com/articles/108433-etiology-diag

**14. Please provide references to scientific papers on guidelines for the tinnitus. Only the bibliographic data of the papers is required.**

1. Langguth, B., Kleinjung, T., Schlee, W., Vanneste, S., & Kreuzer, P. M. (2023). Tinnitus Guidelines and Their Evidence Base. Journal of Clinical Medicine, 12(9), 3087. https://doi.org/10.3390/jcm12093087
2. Tunkel, D. E., Bauer, C. A., Sun, G. H., Rosenfeld, R. M., Chandrasekhar, S. S., Cunningham, E. R., ... & Slattery, W. H. (2014). Clinical practice guideline: tinnitus. Otolaryngology–head and neck surgery, 151(2_suppl), S1-S40. [invalid URL removed]

**15. Please provide references to scientific papers on guidelines for improving voice outcomes after thyroid surgery. Only the bibliographic data of the papers is required.**

1. American Academy of Otolaryngology–Head and Neck Surgery Foundation. Clinical practice guideline: improving voice outcomes after thyroid surgery. Otolaryngol Head Neck Surg. 2013 Jul;149(1):1-37. doi: 10.1177/0194599813487088.
2. Russell JO, Hartman MJ, Clark J, Balakrishnan K, Benninger MS, Berke GS, et al. Clinical Practice Guideline : Improving Voice Outcomes after Thyroid Surgery. Otolaryngol Head Neck Surg. 2021;165:S1-S37.

**16. Please provide references to scientific papers on guidelines for the hoarseness (dysphonia). Only the bibliographic data of the papers is required.**

1. Stachler, R. J., Francis, D. O., Schwartz, S. R., et al. (2018). Clinical Practice Guideline: Hoarseness (Dysphonia) (Update). Otolaryngology–Head and Neck Surgery, 158(1_suppl), S1-S42. doi:10.1177/0194599817750960

**17. Please provide references to scientific papers on guidelines for nutritional management in head and neck cancer. Only the bibliographic data of the papers is required.**

Certainly, here are some references to scientific papers on guidelines for nutritional management in head and neck cancer:

1. Clinical practice guidelines on nutrition management in head andneck cancer: A European Society for Clinical Nutrition and Metabolism (ESPEN) guideline, Clinical Nutrition, Volume 41, Issue 4, 2022, Pages 655-681.
2. Nutrition and head and neck cancer: A review of current evidence, Head & Neck, Volume neck cancer: A review, Nutrition in Clinical Practice, Volume 35, Issue 1, 2020, Pages 3-14.
3. Nutritional support in head and neck cancer: A review, Nutrition, Volume 74, 2020, Pages 110829.

Please note that this is not an exhaustive list and many more articles are available on this topic. You can search for more publications using relevant keywords on scientific databases.

**18. Please provide references to scientific papers on guidelines for the acute otitis externa. Only the bibliographic data of the papers is required.**

Certainly! Here are some references to scientific papers on guidelines for acute otitis externa, providing the bibliographic data as requested:

1. Clinical practice guideline: acute otitis externa

   Rosenfeld RM, Brown L, Cannon CR, Dolor RJ, Ganiats TG, Hannley MT, Kokemueller JE, Marcy SM, Roland PS, Schwartz RH, Shekelle PG, Thomas PA. Otolaryngol Head Neck Surg. 2014 Mar;150(1 Suppl):S1-24. doi: 10.1177/0194599813517294.

2. Management of acute otitis externa: A practical guide

   Kesser BW. Am Fam Physician. 2018 Jan 1;97(1):18-24.

3. Acute otitis externa

   Osguthorpe JD, Nielsen DR. Am Fam Physician. 2006 Oct 1;74(7):1147-52, 1154.

These references should provide you with a good starting point for your research on acute otitis externa guidelines. Please note that this is not an exhaustive list, and there may be other relevant papers available.

Additionally, it's important to consult the latest clinical practice guidelines and recommendations from reputable organizations such as the American Academy of Otolaryngology-Head and Neck Surgery or your local medical societies, as these guidelines are constantly updated based on the latest research and evidence.

**19. Please provide references to scientific papers on guidelines for management of salivary gland tumours. Only the bibliographic data of the papers is required.**

1. Geiger, J. L., Ismaila, N., Caudell, J. J., Chau, N., Glastonbury, C., Y, H., Licitra, L., & Moore, M. G. (2022). Management of Salivary Gland Malignancy: ASCO Guideline. Journal of Clinical Oncology, 40(13), 1493–1513. https://doi.org/10.1200/JCO.21.00449

2. van Herpen, C., Vander Poorten, V., Skalova, A., et al. (2022). Clinical Practice Guideline – Salivary Gland Cancer. European Society for Medical Oncology.

https://www.esmo.org/guidelines/guidelines-by-topic/head-and-neck-cancers/salivary-gland-cancer

3. Williams, M. D., Gupta, R., Lewis, M. A., & Evans, B. T. (2016). Management of Salivary Gland Tumours: United Kingdom National Multidisciplinary Guidelines. Clinical Otolaryngology, 41(S1), 2–6. https://doi.org/10.1111/coa.12592

4. McGurk, M., & Renehan, A. G. (2007). Controversies in the management of salivary gland tumours. The Lancet Oncology, 8(1), 75–84. [invalid URL removed]

**20. Please provide references to scientific papers on guidelines for the peritonsillar abscess. Only the bibliographic data of the papers is required.**

1. Galioto NJ. Peritonsillar Abscess. Am Fam Physician. 2017 May 15;95(10):623-628.

2. Wong DK, Brown J, Lee KC, Ahuja V. Peritonsillar abscess: a 10-year review of 229 cases. Ear Nose Throat J. 2008 Aug;87(8):457-61.

3. Herzon FS, Harris P. Peritonsillar abscess: incidence, current management practices, and a proposal for treatment guidelines. Laryngoscope. 1995 Jul;105(7 Pt 1):728-31.

Źródła

https://www.studocu.com/es-mx/document/universidad-autonoma-de-guadalajara/microbiologia/faringoamigdalitis-bacteriana/40823381

**21. Please provide references to scientific papers on guidelines for the diagnosis of noise-induced hearing loss. Only the bibliographic data of the papers is required.**

1. Moore, B.C.J., Lowe, D.A., Cox, G. (2022). Guidelines for diagnosing and quantifying noise-induced hearing loss. Trends in Hearing, 26, 1-21.

2. Dobie, R. A. (2001). Medical-Legal Evaluation of Hearing Loss. Otolaryngologic Clinics of North America, 34(3), 641-658.

3. Rabinowitz, P. M. (2000). Medicolegal Evaluation of Hearing Loss. American Journal of Industrial Medicine, 37(4), 346-354.

4. Guideline on the diagnosis of noise-induced hearing loss for medicolegal purposes (2000). Scandinavian Audiology. 29: 1, 4-13.

Please note that the Moore et al. (2022) paper has been criticized for its high false-positive rates.

Źródła

https://www.researchgate.net/publication/373664272_Guidelines_for_diagnosis_of_noise-induced_hearing_loss_and_their_specificity

**22. Please provide references to scientific papers on guidelines for the nasal provocations with aspects on nasal patency, airflow, and airflow resistance. Only the bibliographic data of the papers is required.**

1. Clement, P. A. R., & Gordts, F. (1998). Guidelines for nasal provocations with aspects on nasal patency, airflow and airflow resistance. International Committee on Objective Assessment of the Nasal Airways, International Rhinologic Society. Rhinology, 36(2), 56-60.

Źródła

https://med.uth.edu/orl/texas-sinus-institute/research-2/research-2/basic-science-research/

**23. Please provide references to scientific papers on guidelines for the school-based bilingual assessment. Only the bibliographic data of the papers is required.**

1. Caesar, L. G., & Kohler, P. D. (2007). The state of school-based bilingual assessment: Actual practice versus recommended guidelines. Language, Speech, and Hearing Services in Schools, 38(3), 190-200.
2. Bedore, L. M., & Peña, E. D. (2008). Assessment of bilingual children for identification of language impairment: Current findings and implications for practice. International Journal of Bilingual Education and Bilingualism, 11(1), 1-29.

Please note that these are just two examples of scientific papers on guidelines for school-based bilingual assessment. There are many other resources available, including journal articles, book chapters, and technical reports.

Źródła

journals.us.edu.pl/index.php/LOGOPEDIASILESIANA/article/view/8279

dldandme.org/bilingualism-and-dld/

benjamins.com/catalog/tilar.29.int

**24. Please provide references to scientific papers on guidelines for the acquired apraxia of speech. Only the bibliographic data of the papers is required.**

1. Wambaugh, J. L., Duffy, J. R., McNeil, M. R., Robin, D. A., & Ballard, K. J. (2006). Treatment guidelines for acquired apraxia of speech: A synthesis and evaluation of the evidence. Journal of Medical Speech-Language Pathology, 14(4), S341-S354.
2. Wambaugh, J. L., McNeil, M. R., Robin, D. A., Duffy, J. R., & Ballard, K. J. (2006). Treatment guidelines for acquired apraxia of speech: Treatment descriptions and recommendations. Journal of Medical Speech-Language Pathology, 14(4), S355-S368.

Źródła

www.scribd.com/document/130763100/Apraxia-of-Speech

www.scribd.com/document/335522214/Speech-M

**25. Please provide references to scientific papers on guidelines for the safe tracheostomy during the COVID-19 pandemic. Only the bibliographic data of the papers is required.**

Źródła

portal.amelica.org/ameli/journal/351/3514271005/html/

ccforum.biomedcentral.com/articles/10.1186/s13054-021-03674-7

www.cureus.com/articles/127526-outcomes-of-surgical-tracheostomy-on-mechanically-ventilated-covid-19-patients-admitted-to-a-private-tertiary-hospital-in-tanzania

covid19.researcher.life/article/structural-facto
